# REACT-1 round 15 final report: Increased breakthrough SARS-CoV-2 infections among adults who had received two doses of vaccine, but booster doses and first doses in children are providing important protection

**DOI:** 10.1101/2021.12.14.21267806

**Authors:** Marc Chadeau-Hyam, Oliver Eales, Barbara Bodinier, Haowei Wang, David Haw, Matthew Whitaker, Caroline E. Walters, Jakob Jonnerby, Christina Atchison, Peter J. Diggle, Andrew J. Page, Deborah Ashby, Wendy Barclay, Graham Taylor, Graham Cooke, Helen Ward, Ara Darzi, Christl A. Donnelly, Paul Elliott

## Abstract

**Background:** It has been nearly a year since the first vaccinations against SARS-CoV-2 were delivered in England. The third wave of COVID-19 in England began in May 2021 as the Delta variant began to outcompete and largely replace other strains. The REal-time Assessment of Community Transmission-1 (REACT-1) series of community surveys for SARS-CoV-2 infection has provided insights into transmission dynamics since May 2020. Round 15 of the REACT-1 study was carried out from 19 October to 5 November 2021.

**Methods:** We estimated prevalence of SARS-CoV2 infection and used multiple logistic regression to analyse associations between SARS-CoV-2 infection in England and demographic and other risk factors, based on RT-PCR results from self-administered throat and nose swabs in over 100,000 participants. We estimated (single-dose) vaccine effectiveness among children aged 12 to 17 years, and among adults compared swab-positivity in people who had received a third (booster) dose with those who had received two vaccine doses. We used splines to analyse time trends in swab-positivity.

**Results:** During mid-October to early-November 2021, weighted prevalence was 1.57% (1.48%, 1.66%) compared to 0.83% (0.76%, 0.89%) in September 2021 (round 14). Weighted prevalence increased between rounds 14 and 15 across most age groups (including older ages, 65 years and over) and regions, with average reproduction number across rounds of R=1.09 (1.08, 1.11). During round 15, there was a fall in prevalence from a maximum around 20-21 October, with an R of 0.76 (0.70, 0.83), reflecting falls in prevalence at ages 17 years and below and 18 to 54 years. School-aged children had the highest weighted prevalence of infection: 4.95% (4.39%, 5.58%) in those aged 5 to 12 years and 5.21% (4.61%, 5.87%) in those aged 13 to 17 years. In multiple logistic regression, age, sex, key worker status and presence of one or more children in the home were associated with swab positivity. There was evidence of heterogeneity between rounds in swab positivity rates among vaccinated individuals at ages 18 to 64 years, and differences in key demographic and other variables between vaccinated and unvaccinated adults at these ages. Vaccine effectiveness against infection in children was estimated to be 56.2% (41.3%, 67.4%) in rounds 13, 14 and 15 combined, adjusted for demographic factors, with a similar estimate obtained for round 15 only. Among adults we found that those who received a third dose of vaccine were less likely to test positive compared to those who received only two vaccine doses, with adjusted odds ratio (OR) =0.38 (0.26, 0.55).

**Discussion:** Swab-positivity was very high at the start of round 15, reaching a maximum around 20 to 21 October 2021, and then falling through late October with an uncertain trend in the last few days of data collection. The observational nature of survey data and the relatively small proportion of unvaccinated adults call into question the comparability of vaccinated and unvaccinated groups at this relatively late stage in the vaccination programme. However, third vaccine doses for eligible adults and the vaccination of children aged 12 years and over are associated with lower infection risk and, thus, remain a high priority (with possible extension to children aged 5-12 years). These should help reduce SARS-CoV-2 transmission during the winter period when healthcare demands typically rise.

## Introduction

Since May 2020 the REal-time Assessment of Community Transmission-1 (REACT-1) study [1,2] has been tracking the spread of the SARS-CoV-2 virus in England approximately monthly. While the first REACT-1 survey (May 2020) captured the decline of the first wave in England, subsequent surveys have characterised the second and third waves. The third wave of infection began coincident with the rapid spread of the Delta variant of SARS-CoV-2 from May 2021. The third wave generated peaks in the case incidence in mid-July, early September and most recently in mid-October 2021. However, none of these was as high as the peak of the second wave which was observed a week after Christmas 2020.

The national vaccination programme against COVID-19 in England (along with the other countries in the United Kingdom [UK]) began in December 2020, with those at highest risk of exposure (health care workers) and people at highest risk of serious outcomes (those who are older and/or with particular health conditions) being offered the first doses. Over time the groups being offered vaccination were extended to include all adults, then children aged 16 and 17 years, and subsequently children aged 12 to 15 years. Although vaccines are highly successful at reducing serious illness, hospitalisation and death associated with SARS-CoV-2 infection [3,4], individuals who have received two doses of vaccine have a lower, but still appreciable, risk of becoming infected with the Delta variant in the home compared with people who are unvaccinated [5].

The national vaccine programme was extended to school-aged children aged 12 to 15 years (single dose) in September 2021, while third (booster) doses, at least six months (or at least five months for the most vulnerable [6]) following the second dose, are being offered to health and social care workers, all those aged 50 years and over as well as younger people at risk. By mid-November 2021, over 12,000,000 people in the UK had received booster doses. At the same time it was announced that the offer of booster doses would be extended to all those over 40 years of age [7] and that a second dose of the Pfizer-BioNTech vaccine will be offered to 16 and 17 year olds throughout the UK [8].

A study in Israel compared infection prevalence and severe illness among adults aged 60 years and older who had received two vaccine doses with those who had also received a booster dose. At least 12 days after the booster dose, the rate of confirmed infections was 11.3 (95% CI: 10.4, 12.3) times lower and the rate of severe illness was 19.5 (95% CI: 12.9, 29.5) lower in those who had received the booster compared to those who had received only two doses of vaccine [9]. These results are consistent with a detailed immunological study of 23 adults that demonstrated that a booster (third) dose of Pfizer-BioNTech increased SARS-CoV-2 neutralization administered 7 to 9 months after the second dose [10].

Here we present findings from round 15 of REACT-1 showing trends in SARS-CoV-2 prevalence from mid-October to early November 2021 in England, based on self-administered swabs tested by reverse-transcriptase polymerase chain reaction (RT-PCR). In addition to analysing RT-PCR swab-positivity, demographic and other risk factor data obtained from a random sample of the population of England at ages 5 years and over, we assessed the effects of a third vaccine dose on swab-positivity and estimated vaccine effectiveness against infection among children ages 12 to 17 years. The study was carried out against a backdrop of dominance of the Delta variant in England alongside continued rollout of the vaccination programme -- including in children aged 12 years and over -- and rapid implementation of the booster programme.

## Methods

### Study Population

The REACT-1 study methods are available elsewhere [1]. Data collection has taken place each month over a two- to three-week period since May 2020, except for December 2020 and August 2021. The present report relates to data on 100,112 individuals obtained during round 15, which was carried out from 19 October to 5 November 2021 (including a further 93 people who took part from 6 to 8 November 2021); results were compared to those for 100,527 individuals in round 14 (9 to 27 September 2021) [11]. At each round, a random cross-section of the population of England (ages 5 years and over) was invited into the study using as the sampling frame the National Health Service (NHS) list of patients registered with a general practitioner in England, which is held by NHS Digital. Up to round 11 (15 April to 3 May 2021) sampling was designed to achieve approximately equal numbers of participants at the level of lower-tier local authority (LTLA, n=315 in England, with Isles of Scilly combined with Cornwall and the City of London with Westminster). From round 12 (20 May to 7 June 2021) onwards, we invited a random sample of the population in proportion to population size at LTLA level. This had the effect of increasing the numbers sampled in areas of higher population density and reducing the numbers in more sparsely populated areas. However, prevalence estimates should not have been affected (other than precision) as weights are applied to yield estimates that are representative of England as a whole. In addition, from round 14, we altered how swabs (for RT-PCR testing) were collected. Up until round 13 (24 June to 12 July 2021) we used dry swabs which were collected from the participant’s home by courier on a cold chain. We then switched in round 14 to ‘wet’ swabs (transported in saline), which were then, in randomised fashion, either picked up by courier (without cold chain) or sent by priority post, with no difference being detected between the two methods [11]. In the present report (round 15) swabs were only returned using the priority postal service.

### RT-PCR testing

Participants were sent written and video instructions and asked to obtain a self-administered throat and nose swab at home (or their parent/guardian was asked to administer the swab for children aged 12 years and under). Swabs were then sent for RT-PCR testing for SARS-CoV-2, with a positive test being recorded if both N gene and E gene targets were detected or if N gene was detected with cycle threshold (Ct) value below 37.

### Demographic and questionnaire data

We obtained information on age, sex and residential location from the NHS register, and additional information on ethnicity, household size, occupation, past medical history, potential contact with a COVID-19 case, symptoms and other variables via registration and through an online or telephone questionnaire [12].

### Viral genome sequencing

We sent samples that tested positive (with N gene Ct values < 32 and sufficient volume) to the Quadram Institute, Norwich, UK, for viral genome sequencing. The ARTIC protocol [13] (version 4) was used for viral RNA amplification and CoronaHiT for preparation of sequencing libraries [14]. Sequencing data were analysed using the ARTIC bioinformatic pipeline [15] with lineages assigned using PangoLEARN (version 2021-11-4) [16].

### Statistical Analyses

We used R software for the statistical analyses [17]. We calculated unweighted prevalence of swab-positivity as the proportion testing positive on RT-PCR. We then used rim weighting [18] to provide estimates of prevalence that were weighted to be representative of the population of England as a whole. We used multiple logistic regression to estimate odds ratios for the effects of demographic and other variables on swab-positivity, both age-and-sex adjusted and mutually adjusted for the other variables considered.

We used an exponential model of growth or decay to investigate temporal trends in swab-positivity, assuming that numbers of positive samples out of the total number of samples per day arose from a binomial distribution. Swabs were assigned to day of swabbing where reported or to day of first scan of the sample by the Post Office otherwise (samples were excluded from the temporal analyses when neither swab date nor scan were recorded). We estimated posterior credible intervals using a bivariate No-U-Turn Sampler with a uniform prior distribution for the probability of swab-positivity on day of swabbing and the growth rate [19]. To estimate the reproduction number R, we assumed a gamma-distributed generation time with shape parameter, n=2.29 and rate parameter *β*=0.36 (corresponding to a mean generation time of 6.29 days) [20].

We fit a Bayesian penalised-spline (P-spline) model [21] to the daily data using a No-U-Turn Sampler in logit space to visualise trends in swab-positivity over time. The data were partitioned into approximately 5-day sections by regularly spaced knots, adding further knots beyond the study period to minimise edge effects. We also fit P-splines separately to three broad age groups (17 years and under, 18 to 54 years, 55 years and over) using a smoothing parameter obtained from the model fit to all data. We then examined the link between swab-positivity data in REACT-1 and publicly available hospitalisations and COVID-19 mortality data (deaths within 28 days of a positive test). First, we fit an analogous P-spline model to both sets of publicly available data assuming a negative-binomial likelihood, with an extra overdispersion parameter that was assumed to have a non-informative constant prior distribution. We then selected a random sample (N=1,000) from the posterior distributions of the P-spline model fits and fit a simple two-parameter model to the first seven rounds of REACT-1 daily swab-positivity data (seven rounds were selected to represent the pre-vaccination period). The two parameters were a discrete time-lag between swab-positivity and hospitalisation/death time series (1,000 time series) and a population-adjusted scaling factor that corresponds to the probability of those testing swab-positive on day i being hospitalised/dying on day i+**τ**, where **τ** is the time-lag parameter.

We estimated the daily growth rate of the proportion of AY.4.2 Delta sub-lineage relative to all other lineages using a Bayesian logistic regression model and assuming a binomial likelihood for the daily proportion of AY.4.2.

We estimated smoothed LTLA prevalence of swab-positivity using a ‘nearest-neighbour’ approach. From a subsample of individuals (N=15) per LTLA we obtained the median number of nearest neighbours (within a 30km radius) of each individual, calculated neighbourhood prevalence per individual, then obtained the mean neighbourhood prevalence across all N=15 individuals per LTLA. We defined individual neighbourhood prevalence as the number of nearest neighbours with positive tests divided by the total number of nearest neighbours.

To investigate the effect of a third vaccine dose, we used multiple logistic regression to estimate (round 15) odds ratios of swab-positivity comparing adults who had received a third dose of vaccine to those having had two doses. Estimates were made unadjusted and with adjustments for age and sex, and additionally for Index of Multiple Deprivation (IMD), region, and ethnicity.

We estimated vaccine effectiveness against infection among children ages 12 to 17 years by combining data from round 13 to 15 (to increase statistical power) and for round 15 alone. We used data linked (with consent) to the national COVID-19 vaccination programme to obtain information on who had received a vaccination. Using the dates from the data linkage, a child was considered to have been vaccinated (one dose) 14 days after administration of the vaccine (being considered unvaccinated before then). We estimated vaccine effectiveness as 1 - odds ratio (OR), where we estimated OR from a logistic regression model comparing swab positivity among vaccinated and unvaccinated individuals, with adjustment for round, then sequentially round, age and sex, and additionally IMD, region and ethnicity.

## Results

A total of 859,184 participants were invited to participate in round 15, 143,193 (16.7%) registered, of whom 100,112 (69.9%) provided a swab with a valid result from RT-PCR (Supplementary Table 1).

### COVID-19 prevalence in England 19 October - 5 November 2021

Of the 100,112 valid swabs, 1,399 were positive yielding a weighted prevalence of 1.57% (1.48%, 1.66%), nearly two-fold higher than that estimated in round 14 at 0.83% (0.76%, 0.89%). We observed an increase in weighted prevalence between round 14 and round 15 across most ages (including those aged 65 years and over) and regions, with average reproduction number across rounds of R=1.09 (1.08, 1.11). Increases were also seen by occupation, ethnicity (Supplementary Table 2a), lifestyle (household size, number of children in the household), medical (symptoms reported in the month prior to swabbing) and social (contact with COVID cases and deprivation) factors (Supplementary Table 2b). We observed the highest weighted prevalence in those aged 13 to 17 years at 5.21% (4.61%, 5.87%) and those aged 5 to 12 years at 4.95% (4.39%, 5.58%) (Supplementary Table 2a, Supplementary Figure 1-A).

The highest weighted prevalence in round 15 by region was observed in South West at 1.97% (1.69%, 2.29%) increasing more than three-fold from round 14 at 0.59% (0.43%, 0.80%) (Supplementary Table 2a, Supplementary Figure 1-B). At LTLA level, the ten highest smoothed estimates of prevalence based on a nearest neighbour method were all found in parts of the South West (Supplementary Figure 2).

We also note that higher weighted prevalence was observed in (i) larger households including five people at 3.12% (2.66%, 3.66%) or six or more people at 2.95% (2.27%, 3.82%) compared to 0.78% (0.65%, 0.94%) in single-person households; (ii) households with one or more children at 2.62% (2.41%, 2.85%) compared to 0.74% (0.67%, 0.81%) in households without children; (iii) in those reporting having been in contact with a confirmed COVID-19 case at 9.13% (8.35%, 9.96%) compared to 0.83% (0.75%, 0.90%) for those without such contact, and (iv) in those reporting classic COVID-19 symptoms in the month prior to swabbing at 7.84% (7.22%, 8.51%) compared to 0.67% (0.60%, 0.75%) in those without symptoms (Supplementary Table 2b).

Multiple logistic models for swab-positivity (Table 1) indicated that after adjusting for all variables, other essential/key workers were at greater risk of swab-positivity than other workers with OR of 1.23 (1.05, 1.44). Mutually adjusted OR for households with one or more children was 1.97 (1.60, 2.42) in round 15 compared to households without children, while increased risk for larger vs smaller households was not observed after adjustment for other covariates (including the number of children in the household).

**Table 1.**
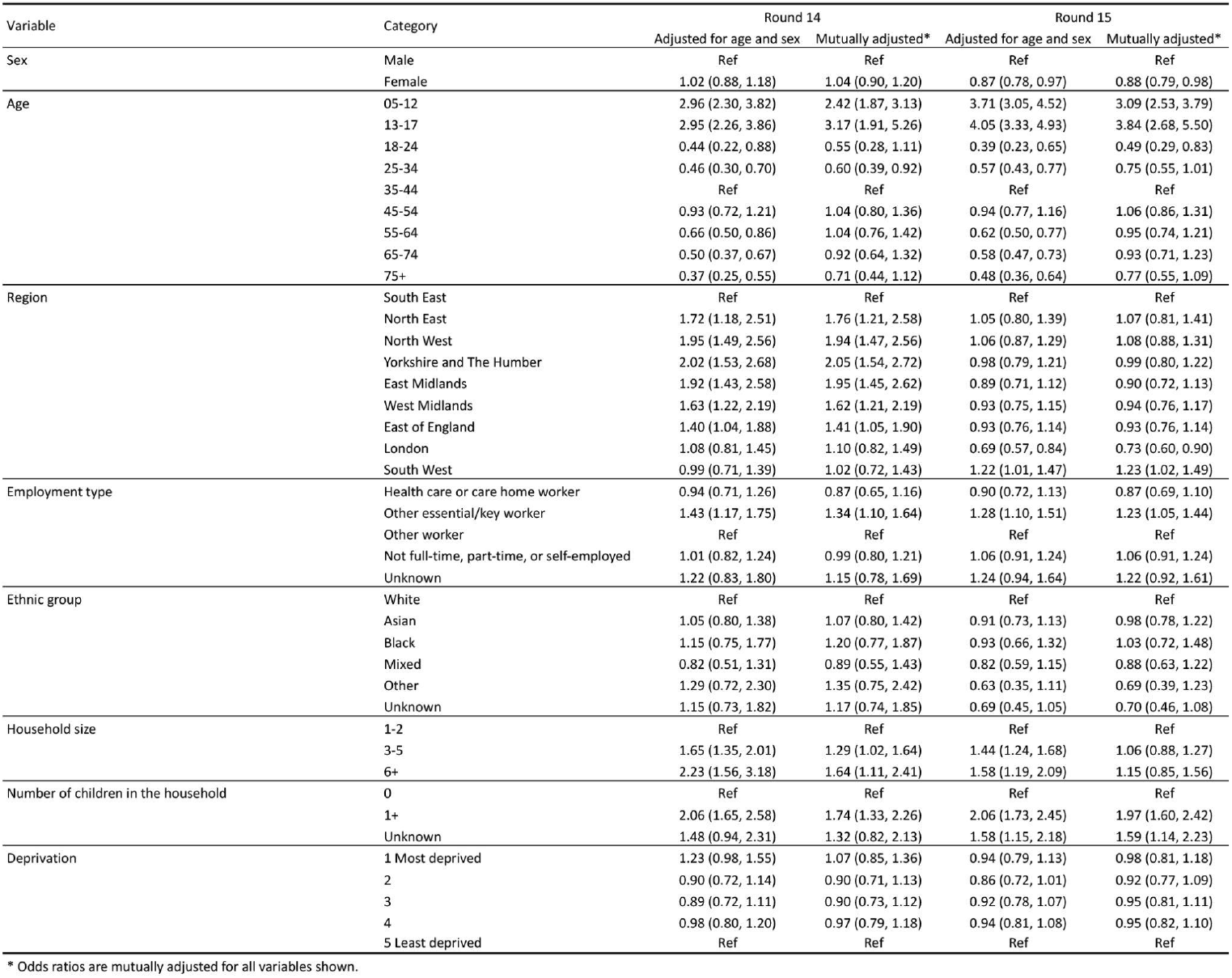
Multiple logistic regression for swab positivity in rounds 14 and 15. Results are presented as Odds Ratios (95% confidence interval) adjusted for age and sex and additionally for all other variables (mutually adjusted OR).

| Variable | Category | Round 14 |  | Round 15 |  |
| --- | --- | --- | --- | --- | --- |
|  |  | Adjusted for age and sex | Mutually adjusted* | Adjusted for age and sex | Mutually adjusted* |
| Sex | Male | Ref | Ref | Ref | Ref |
|  | Female | 1.02 (0.88, 1.18) | 1.04 (0.90, 1.20) | 0.87 (0.78, 0.97) | 0.88 (0.79, 0.98) |
| Age | 05-12 | 2.96 (2.30, 3.82) | 2.42 (1.87, 3.13) | 3.71 (3.05, 4.52) | 3.09 (2.53, 3.79) |
|  | 13-17 | 2.95 (2.26, 3.86) | 3.17 (1.91, 5.26) | 4.05 (3.33, 4.93) | 3.84 (2.68, 5.50) |
|  | 18-24 | 0.44 (0.22, 0.88) | 0.55 (0.28, 1.11) | 0.39 (0.23, 0.65) | 0.49 (0.29, 0.83) |
|  | 25-34 | 0.46 (0.30, 0.70) | 0.60 (0.39, 0.92) | 0.57 (0.43, 0.77) | 0.75 (0.55, 1.01) |
|  | 35-44 | Ref | Ref | Ref | Ref |
|  | 45-54 | 0.93 (0.72, 1.21) | 1.04 (0.80, 1.36) | 0.94 (0.77, 1.16) | 1.06 (0.86, 1.31) |
|  | 55-64 | 0.66 (0.50, 0.86) | 1.04 (0.76, 1.42) | 0.62 (0.50, 0.77) | 0.95 (0.74, 1.21) |
|  | 65-74 | 0.50 (0.37, 0.67) | 0.92 (0.64, 1.32) | 0.58 (0.47, 0.73) | 0.93 (0.71, 1.23) |
|  | 75+ | 0.37 (0.25, 0.55) | 0.71 (0.44, 1.12) | 0.48 (0.36, 0.64) | 0.77 (0.55, 1.09) |
| Region | South East | Ref | Ref | Ref | Ref |
|  | North East | 1.72 (1.18, 2.51) | 1.76 (1.21, 2.58) | 1.05 (0.80, 1.39) | 1.07 (0.81, 1.41) |
|  | North West | 1.95 (1.49, 2.56) | 1.94 (1.47, 2.56) | 1.06 (0.87, 1.29) | 1.08 (0.88, 1.31) |
|  | Yorkshire and The Humber | 2.02 (1.53, 2.68) | 2.05 (1.54, 2.72) | 0.98 (0.79, 1.21) | 0.99 (0.80, 1.22) |
|  | East Midlands | 1.92 (1.43, 2.58) | 1.95 (1.45, 2.62) | 0.89 (0.71, 1.12) | 0.90 (0.72, 1.13) |
|  | West Midlands | 1.63 (1.22, 2.19) | 1.62 (1.21, 2.19) | 0.93 (0.75, 1.15) | 0.94 (0.76, 1.17) |
|  | East of England | 1.40 (1.04, 1.88) | 1.41 (1.05, 1.90) | 0.93 (0.76, 1.14) | 0.93 (0.76, 1.14) |
|  | London | 1.08 (0.81, 1.45) | 1.10 (0.82, 1.49) | 0.69 (0.57, 0.84) | 0.73 (0.60, 0.90) |
|  | South West | 0.99 (0.71, 1.39) | 1.02 (0.72, 1.43) | 1.22 (1.01, 1.47) | 1.23 (1.02, 1.49) |
| Employment type | Health care or care home worker | 0.94 (0.71, 1.26) | 0.87 (0.65, 1.16) | 0.90 (0.72, 1.13) | 0.87 (0.69, 1.10) |
|  | Other essential/key worker | 1.43 (1.17, 1.75) | 1.34 (1.10, 1.64) | 1.28 (1.10, 1.51) | 1.23 (1.05, 1.44) |
|  | Other worker | Ref | Ref | Ref | Ref |
|  | Not full-time, part-time, or self-employed | 1.01 (0.82, 1.24) | 0.99 (0.80, 1.21) | 1.06 (0.91, 1.24) | 1.06 (0.91, 1.24) |
|  | Unknown | 1.22 (0.83, 1.80) | 1.15 (0.78, 1.69) | 1.24 (0.94, 1.64) | 1.22 (0.92, 1.61) |
| Ethnic group | White | Ref | Ref | Ref | Ref |
|  | Asian | 1.05 (0.80, 1.38) | 1.07 (0.80, 1.42) | 0.91 (0.73, 1.13) | 0.98 (0.78, 1.22) |
|  | Black | 1.15 (0.75, 1.77) | 1.20 (0.77, 1.87) | 0.93 (0.66, 1.32) | 1.03 (0.72, 1.48) |
|  | Mixed | 0.82 (0.51, 1.31) | 0.89 (0.55, 1.43) | 0.82 (0.59, 1.15) | 0.88 (0.63, 1.22) |
|  | Other | 1.29 (0.72, 2.30) | 1.35 (0.75, 2.42) | 0.63 (0.35, 1.11) | 0.69 (0.39, 1.23) |
|  | Unknown | 1.15 (0.73, 1.82) | 1.17 (0.74, 1.85) | 0.69 (0.45, 1.05) | 0.70 (0.46, 1.08) |
| Household size | 1-2 | Ref | Ref | Ref | Ref |
|  | 3-5 | 1.65 (1.35, 2.01) | 1.29 (1.02, 1.64) | 1.44 (1.24, 1.68) | 1.06 (0.88, 1.27) |
|  | 6+ | 2.23 (1.56, 3.18) | 1.64 (1.11, 2.41) | 1.58 (1.19, 2.09) | 1.15 (0.85, 1.56) |
| Number of children in the household | 0 | Ref | Ref | Ref | Ref |
|  | 1+ | 2.06 (1.65, 2.58) | 1.74 (1.33, 2.26) | 2.06 (1.73, 2.45) | 1.97 (1.60, 2.42) |
|  | Unknown | 1.48 (0.94, 2.31) | 1.32 (0.82, 2.13) | 1.58 (1.15, 2.18) | 1.59 (1.14, 2.23) |
| Deprivation | 1 Most deprived | 1.23 (0.98, 1.55) | 1.07 (0.85, 1.36) | 0.94 (0.79, 1.13) | 0.98 (0.81, 1.18) |
|  | 2 | 0.90 (0.72, 1.14) | 0.90 (0.71, 1.13) | 0.86 (0.72, 1.01) | 0.92 (0.77, 1.09) |
|  | 3 | 0.89 (0.72, 1.11) | 0.90 (0.73, 1.12) | 0.92 (0.78, 1.07) | 0.95 (0.81, 1.11) |
|  | 4 | 0.98 (0.80, 1.20) | 0.97 (0.79, 1.18) | 0.94 (0.81, 1.08) | 0.95 (0.82, 1.10) |
|  | 5 Least deprived | Ref | Ref | Ref | Ref |

\* Odds ratios are mutually adjusted for all variables shown.

### Epidemic dynamics

P-spline models fit to data from all rounds of REACT-1 (Figure 1-A) showed an increasing weighted prevalence of swab-positivity from round 14 to round 15 followed by a fall during round 15 (Figure 1-B). The posterior distribution of the estimated epidemic curve suggested that the peak in weighted prevalence was reached on around 20 to 21 October (with 95% credible intervals ranging from 15 to 23 October) followed by a fall (Supplementary Figure 3). Our estimates did not exclude an upturn in the last days of round 15 (from 2 November onwards) but this was based on fewer observations in the first week of November compared to the previous days (Figure 1-C). Similar trends of rising prevalence between round 14 and round 15 followed by a fall within round 15 were observed for children aged 17 years and under and adults aged 18 to 54 years but a fall was not observed at ages 55 years and over (Figure 1-D).

**Figure 1.**
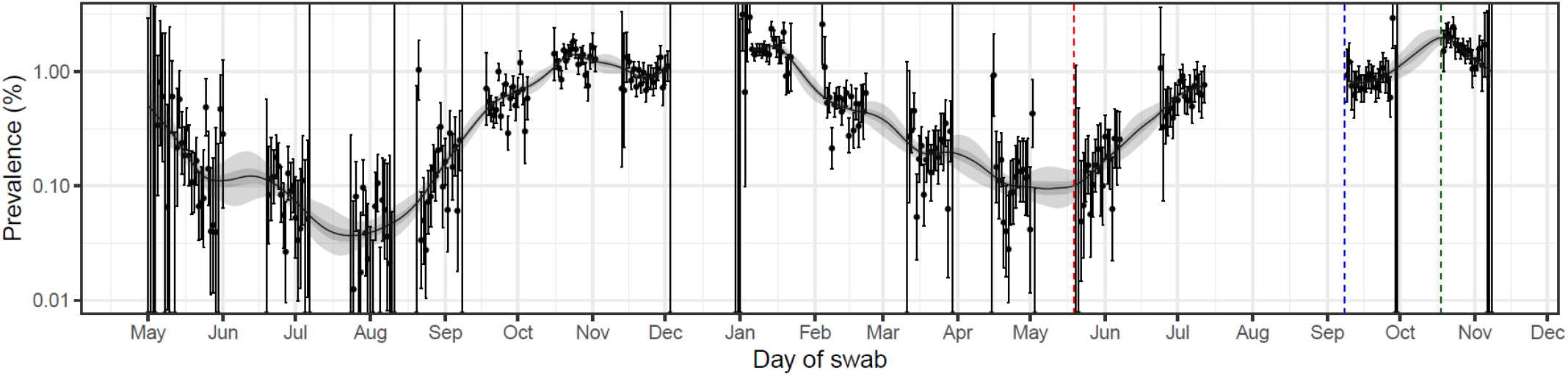
**(A)**. Daily weighted swab-positivity for all 15 rounds of the REACT-1 study (black points with 95% confidence intervals, left-hand y-axis) with P-spline estimates for swab-positivity (solid black line, shaded area is 95% credible interval). Changes in testing procedures are identified by vertical dashed lines. Geographic sampling procedure changed for rounds 12 onwards (red line), round 14 had half of respondents’ swab tests collected by courier and the other half post their swab test (blue line) and for round 15 all respondents posted their swab test (green line).

**Figure 1 B and C.**
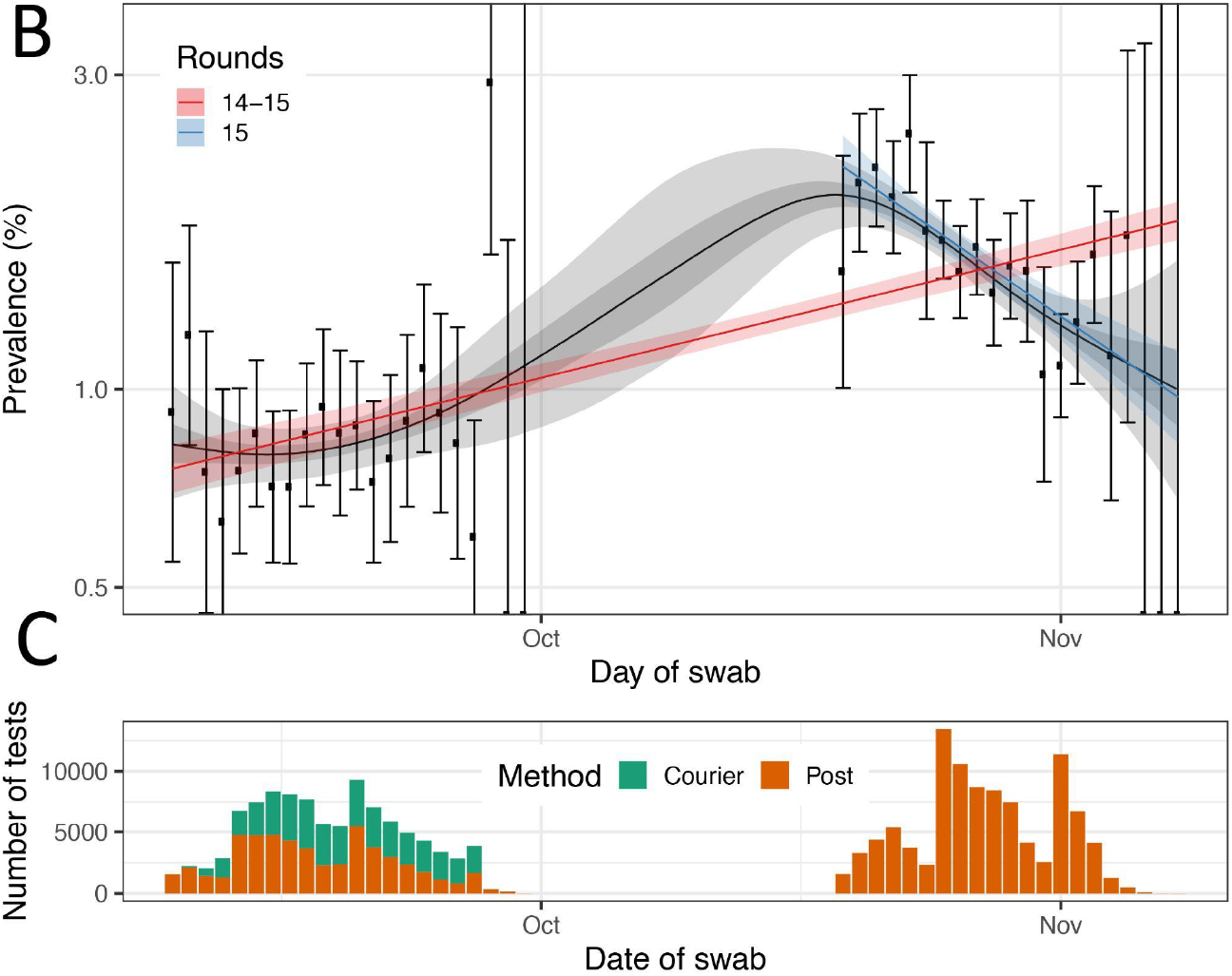
**(B)** Comparison of an exponential model fit to round 14-15 (red) and round 15 only (blue) and a P-spline model fit to all rounds of REACT-1 (black, shown here only for round 14 and 15). Shaded red and blue regions show the 95% posterior credible intervals for the exponential model, and the shaded grey region shows 50% (dark grey) and 95% (light grey) posterior credible interval for the P-spline model. Results are presented for each day (X axis) of sampling for round 14 and round 15 and the prevalence of infection is shown (Y axis) on a log scale. Weighted observations (black dots) and 95% confidence intervals (vertical lines) are also shown. **(C)** Number of samples processed per day during round 14 and round 15. In round 14 the samples shipped by post are represented in orange, and those shipped by courier, in green.

**Figure 1 D.**
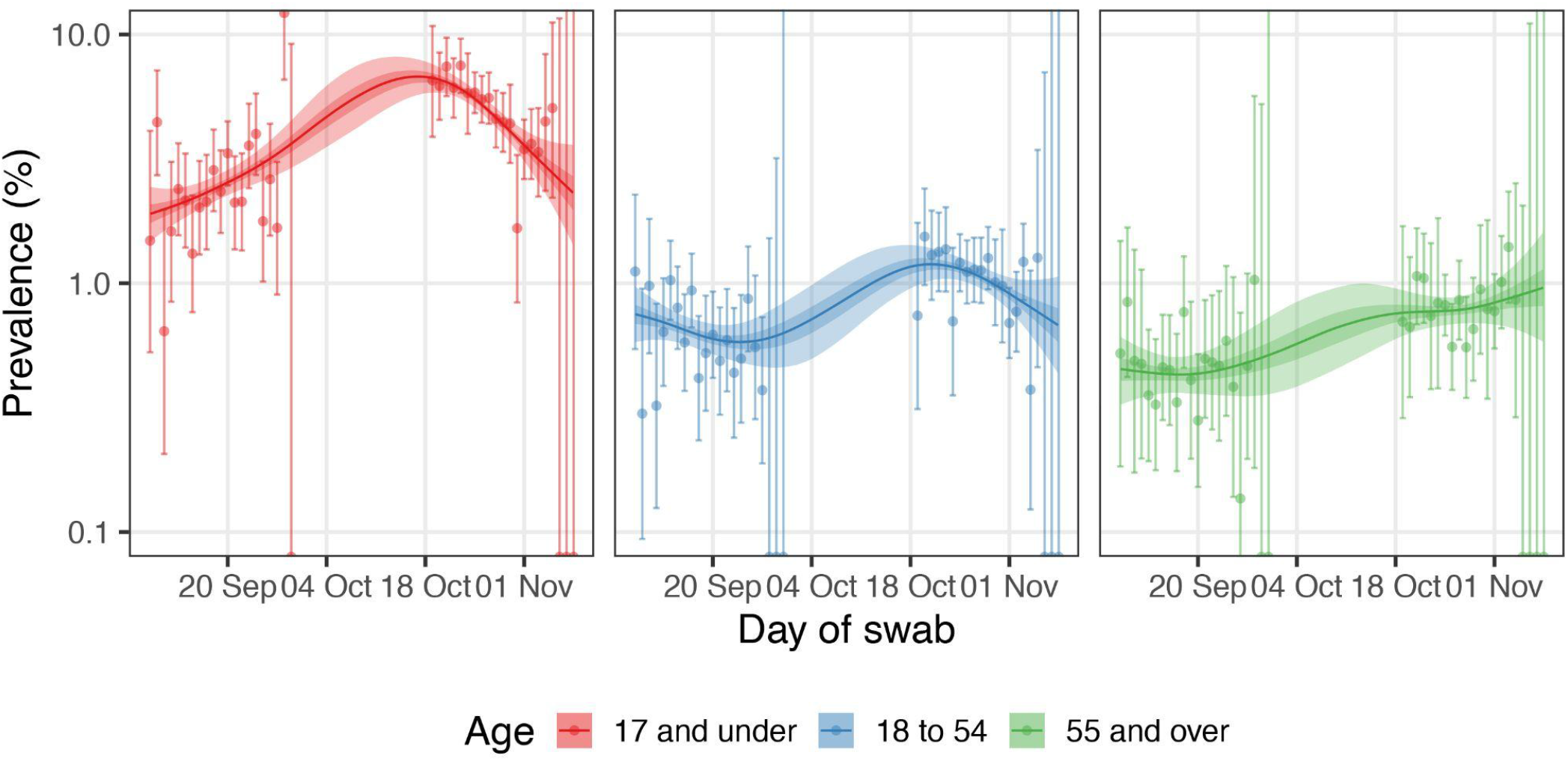
Comparison of P-spline models fit to all rounds of REACT-1 for those aged 17 years and under (red), those aged 18 to 54 years inclusive (blue) and those aged 55 years and over (green). Shown here only for the period of round 14 and round 15. Shaded regions show 50% (dark shade) and 95% (light shade) posterior credible interval for the P-spline models. Results are presented for each day (X axis) of sampling for round 14 and round 15 and the prevalence of swab-positivity is shown (Y axis) on a log scale. Weighted observations (dots) and 95% confidence intervals (vertical lines) are also shown.

An exponential model fit to data from round 15 indicated a fall in weighted prevalence during October 2021 with R=0.76 (0.70, 0.83) and posterior probability that R>1 lower than 0.01. Decreasing weighted prevalence within round 15 was also detected with posterior probability that R>1 below 0.01 at ages 17 years and under and 18 to 54 years, and in East Midlands, South East and South West (Table 2).

**Table 2.**
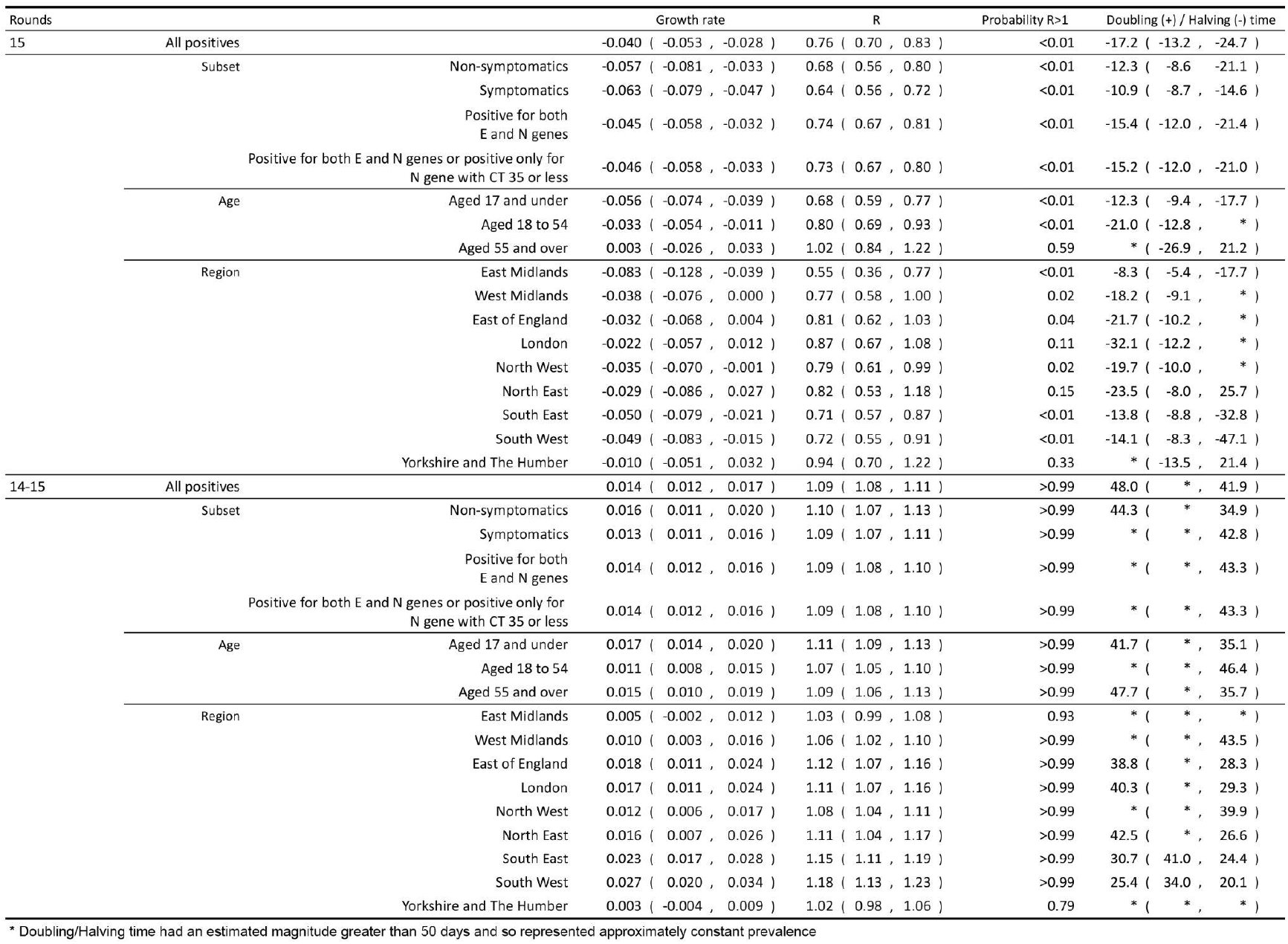
Table of growth rates, reproduction numbers and doubling/halving times from exponential model fits on data from round 15 only (top table) and round 14 and round 15 (bottom table)

### Vaccination and third dose in adults, and single dose in children

Across rounds 13, 14 and 15 (predominantly Delta), vaccination data were obtained (with consent) from linkage to NHS data in 259,936 participants aged 18 to 64 years. Of these, 132,333 had received two vaccine doses at least 14 days prior to swabbing, and 5,025 were unvaccinated across rounds 13, 14 and 15 (Table 3). The number of unvaccinated adults decreased from 2,951 (7.7%) in round 13 to 1,039 (2.0%) in round 14 and 1,035 (2.1%) in round 15, indicating high vaccine uptake at these ages (Figure 2-A). Using a logistic model for vaccination status, we identified key demographic and other differences between the unvaccinated and vaccinated individuals, especially during round 14 and 15 (Figure 2-A) and found evidence of heterogeneity between rounds in swab positivity rates among vaccinated individuals at ages 18 to 64 years. We therefore examined prevalence of swab-positivity by round and vaccination status (Table 3). Unweighted prevalence of infection in unvaccinated adults decreased from 1.49% (1.09%, 2.00%) in round 13 to 1.16% (0.60%, 2.02%) in round 15, while unweighted prevalence of breakthrough infections in those who had received two doses of vaccine more than doubled between round 13 and round 15 from 0.41% (0.35%,0.48%) to 1.10% (1.01%, 1.20%), respectively.

**Table 3.**
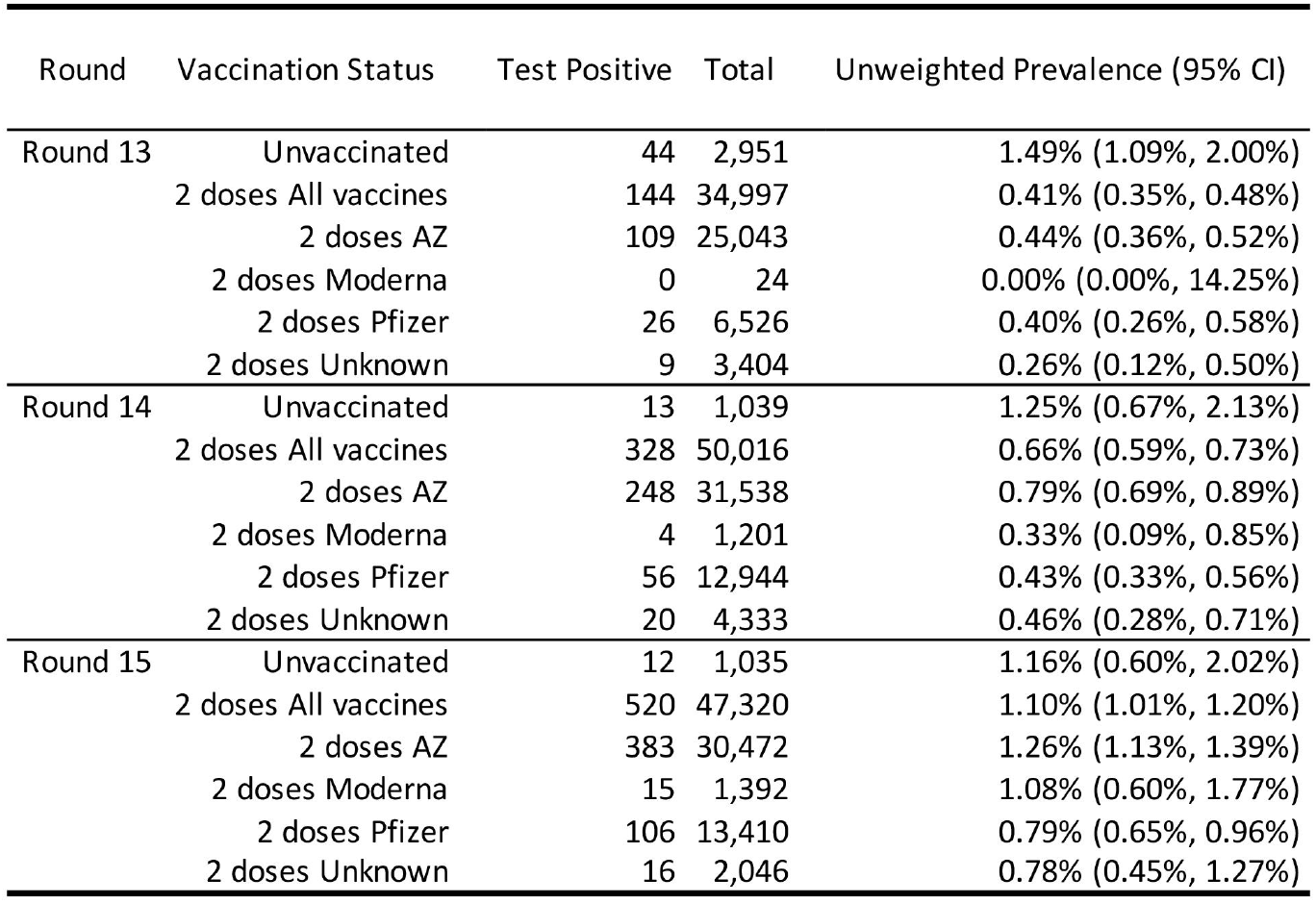
Unweighted prevalence of swab-positivity by vaccination status^1^ and round, for rounds 13, 14 and 15 of REACT-1. Results are presented for linked vaccine status data, for participants aged 18 to 64 years.

| Round | Vaccination Status | Test Positive | Total | Unweighted Prevalence (95% CI) |
| --- | --- | --- | --- | --- |
| Round 13 | Unvaccinated | 44 | 2,951 | 1.49% (1.09%, 2.00%) |
|  | 2 doses All vaccines | 144 | 34,997 | 0.41% (0.35%, 0.48%) |
|  | 2 doses AZ | 109 | 25,043 | 0.44% (0.36%, 0.52%) |
|  | 2 doses Moderna | 0 | 24 | 0.00% (0.00%, 14.25%) |
|  | 2 doses Pfizer | 26 | 6,526 | 0.40% (0.26%, 0.58%) |
|  | 2 doses Unknown | 9 | 3,404 | 0.26% (0.12%, 0.50%) |
| Round 14 | Unvaccinated | 13 | 1,039 | 1.25% (0.67%, 2.13%) |
|  | 2 doses All vaccines | 328 | 50,016 | 0.66% (0.59%, 0.73%) |
|  | 2 doses AZ | 248 | 31,538 | 0.79% (0.69%, 0.89%) |
|  | 2 doses Moderna | 4 | 1,201 | 0.33% (0.09%, 0.85%) |
|  | 2 doses Pfizer | 56 | 12,944 | 0.43% (0.33%, 0.56%) |
|  | 2 doses Unknown | 20 | 4,333 | 0.46% (0.28%, 0.71%) |
| Round 15 | Unvaccinated | 12 | 1,035 | 1.16% (0.60%, 2.02%) |
|  | 2 doses All vaccines | 520 | 47,320 | 1.10% (1.01%, 1.20%) |
|  | 2 doses AZ | 383 | 30,472 | 1.26% (1.13%, 1.39%) |
|  | 2 doses Moderna | 15 | 1,392 | 1.08% (0.60%, 1.77%) |
|  | 2 doses Pfizer | 106 | 13,410 | 0.79% (0.65%, 0.96%) |
|  | 2 doses Unknown | 16 | 2,046 | 0.78% (0.45%, 1.27%) |

<sup>1</sup> Vaccination status for linked data was defined using time since last vaccination. Unvaccinated are those not having received any vaccine dose or one dose less than 14 days before swabbing; double dose vaccinated are those having received their second dose 14 days or more before swabbing

**Figure 2.**
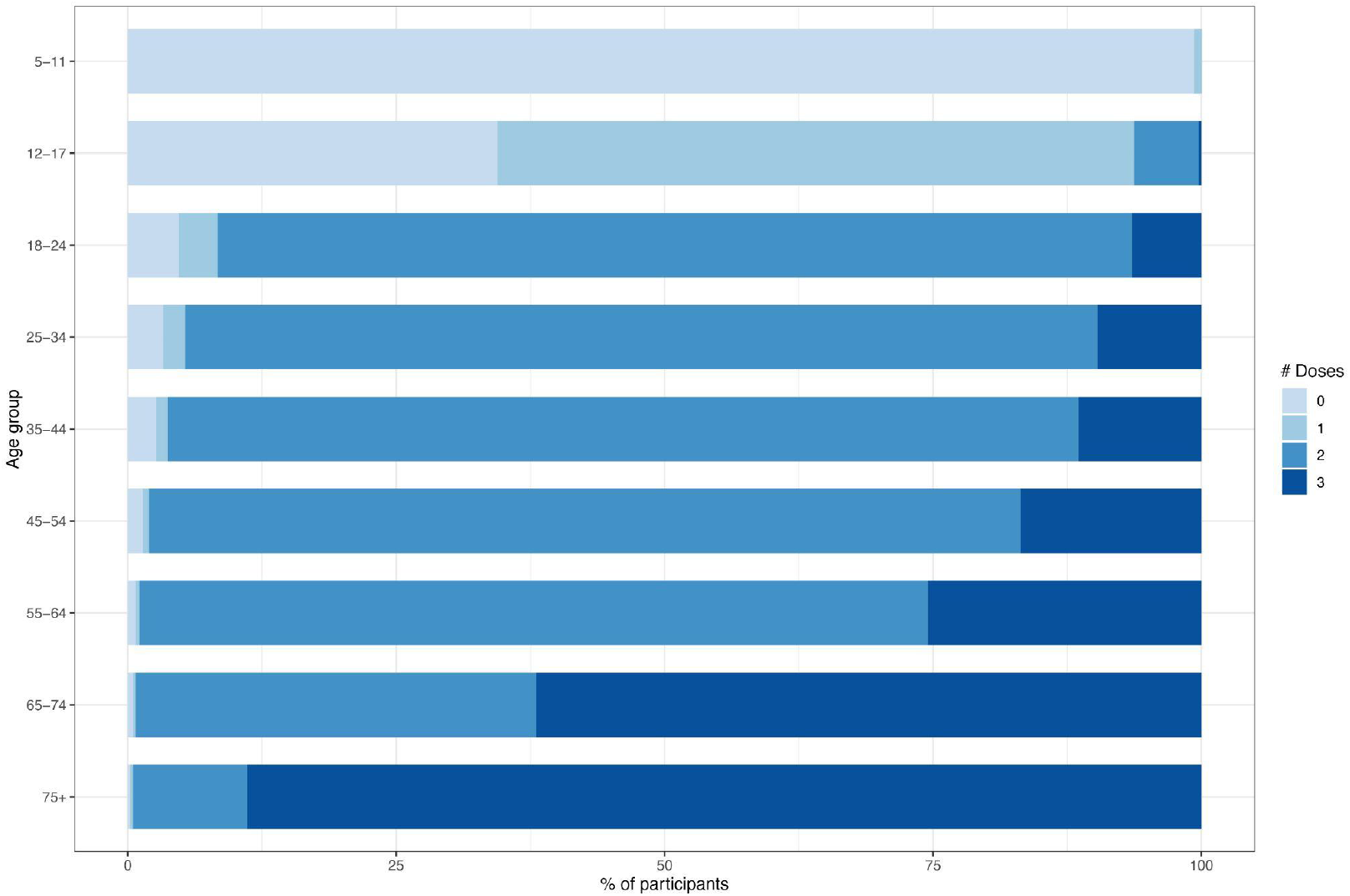
**A**. Distribution of number of vaccine doses received by age group. Results are on linked vaccination data in consenting participants (children aged 12 years were combined with those aged 13 to 17 years, as they were eligible for vaccination).

**Figure 2 B and C.**
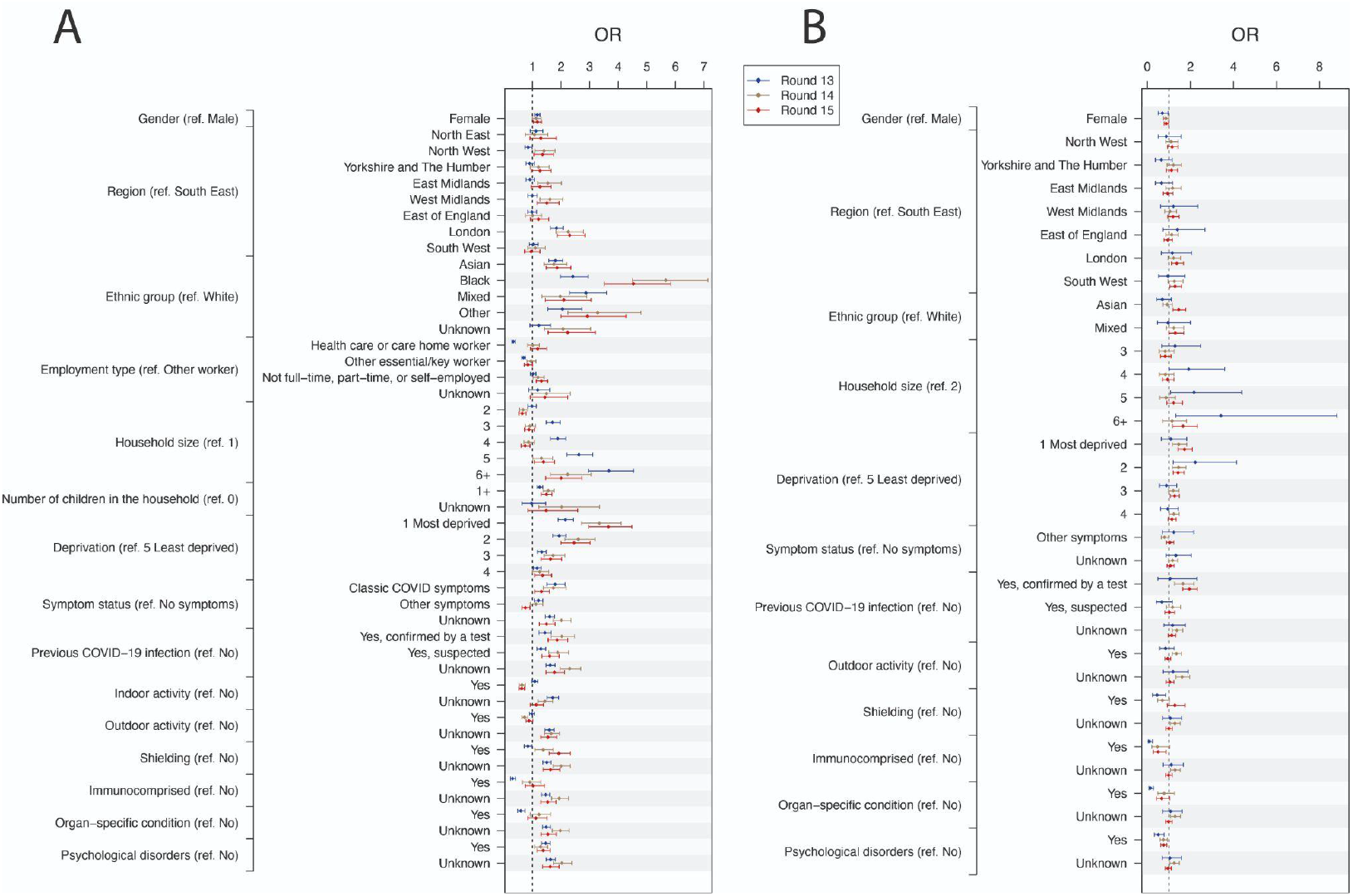
Comparison of the characteristics of the unvaccinated and vaccinated participants of REACT-1 rounds 13-15. For each variable, we present the point estimate and 95% confidence interval of the Odds Ratio (OR) from the logistic model for vaccination status. This is parameterised such that ORs greater than 1 indicate a greater probability of being unvaccinated. Results are presented for round 13 (blue), round 14 (beige), and round 15 (red) in participants aged 18 to 64 years (A), and 12 to 17 years (B). For the 18 to 64 years age group, the model compares double vaccinated to unvaccinated people, and for the 12 to 17 years group, single-or-double vaccinated to unvaccinated children.

In contrast, we found that the (N=11,740) unvaccinated and the (N=3,234) vaccinated children aged 12 to 17 years were broadly comparable (Figure 2-B). However, we found that immunosuppressed children and those suffering from psychological disorders or organ-specific conditions, particularly in round 13, were more likely to have been vaccinated. Using data from rounds 13, 14 and 15, we estimated an unadjusted vaccine effectiveness for a single dose of Pfizer-BioNTech (after 14 days) in children aged 12 to 17 years to be 53.6% (39.5%, 64.5%) (Table 4). With adjustment for age and sex, vaccine effectiveness was estimated at 54.6% (39.3%, 66.0%) and with additional adjustment for index of multiple deprivation, region and ethnicity, it was 56.2% (41.3%, 67.4%). Higher estimates (fully adjusted) were found in those reporting symptoms at 67.5% (51.0%,78.5%). Vaccine effectiveness estimates were similar when children who had received two doses of vaccine were included.

**Table 4.**
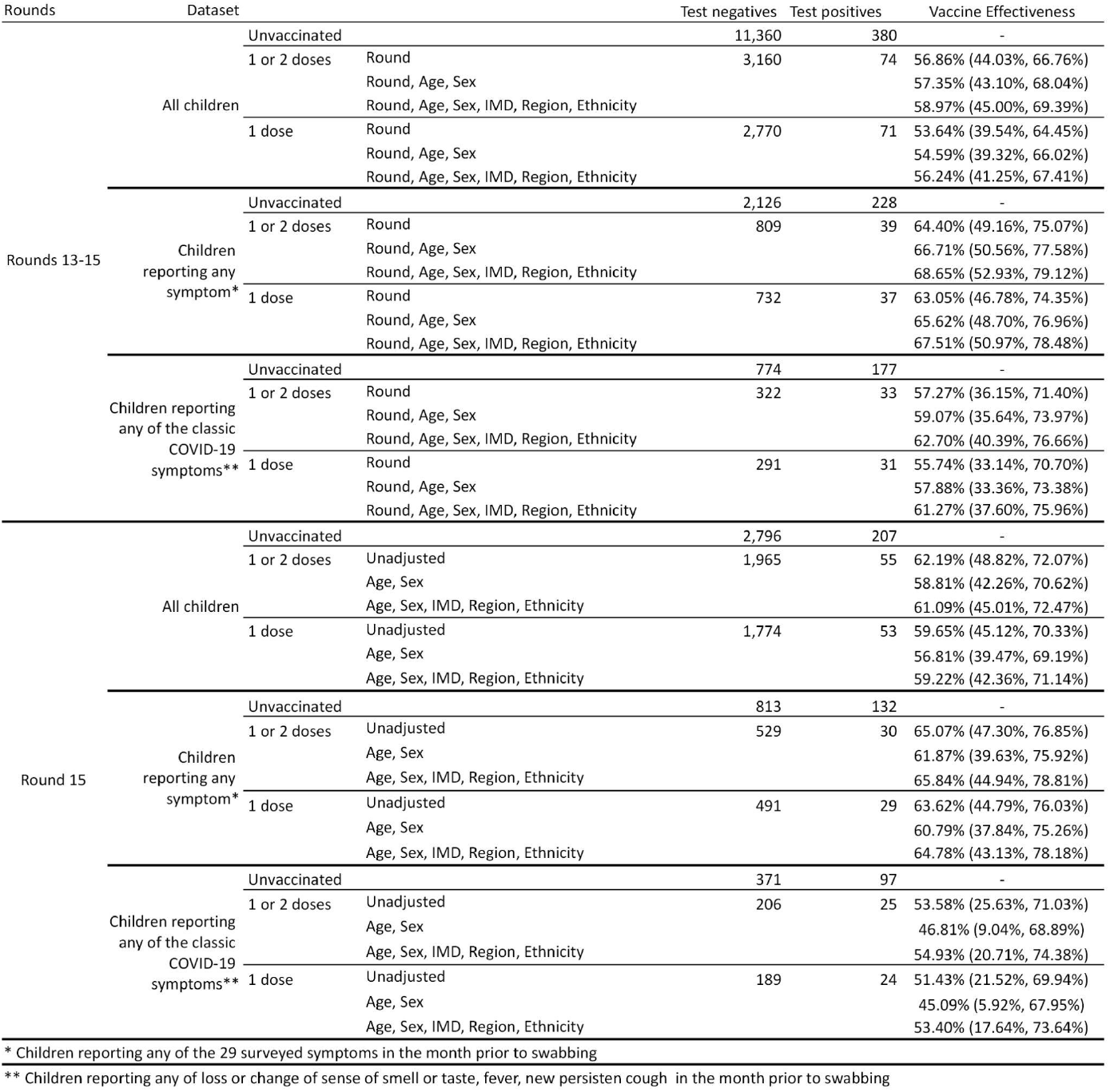
Estimates of vaccine effectiveness against infection for children aged 12 to 17 years in (i) round 13 to round 15, and (ii) round 15 of REACT-1. Estimates are based on a logistic model of swab positivity in (i) children having received one or two vaccine doses and (ii) children having received a single vaccine dose compared to unvaccinated children, for all children and symptomatic children only. Results from round 15 only are unadjusted and additionally adjusted for age and sex, and for Index of Multiple Deprivation (IMD, region, and ethnicity. For the model based on data from rounds 13, 14 and 15, estimates are further adjusted for round.

The fully adjusted OR of swab-positivity associated with a third (booster) dose compared to two doses of vaccine was estimated at OR=0.38 (0.26, 0.55) for all ages and all vaccines combined (Table 5). Similar results were obtained looking at adults aged 50 years and over together with health care workers and care home workers under 50 years of age, with a suggestion that protection was greater if the third dose followed two doses of Pfizer-BioNTech than two doses of AstraZeneca.

**Table 5.**
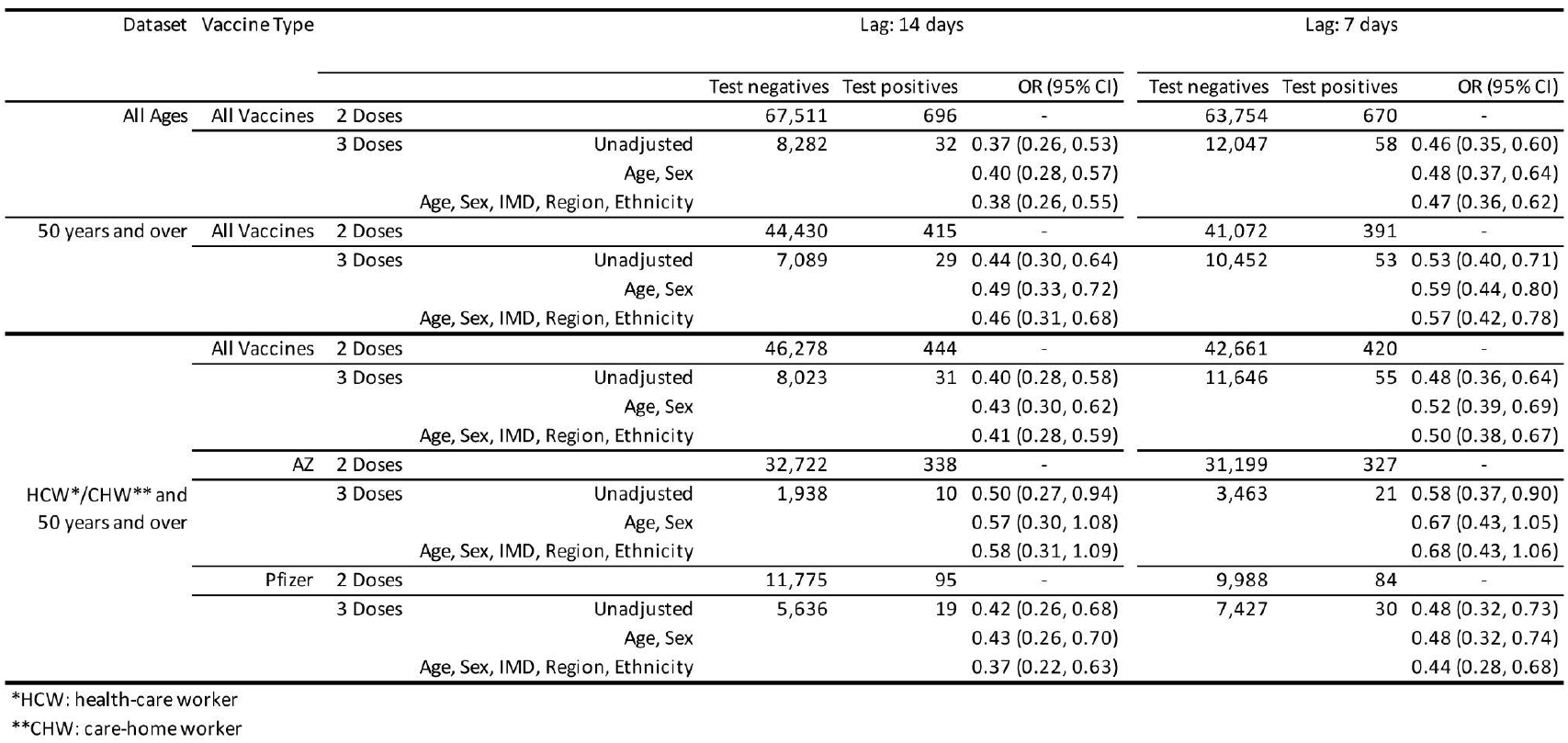
Estimates of the effect of a third vaccine dose on the risk of infection in round 15 participants of REACT-1 having received at least two vaccine doses. Estimates are obtained comparing swab positivity in those having received two vaccine doses and those having received three vaccine doses^2^. Results are presented for the entire round 15 population, for those aged 50 years and over, anf for those either aged 50 years and over or health care or care home workers. In the latter group estimates are given for all vaccines combined and for AZ and Pfizer-BioNTech separately. Odds Ratios (ORs) are shown unadjusted, adjusted for age and sex, and additionally adjusted for Index of Multiple Deprivation (IMD), region, and ethnicity.

### Temporal analyses

We estimated shifting and scaling parameters to overlay daily hospitalisations (Figure 3-A) and, separately, COVID-19 deaths (deaths within 28 days of a positive test, Figure 3-B) onto the daily percentage of people who tested swab-positive in REACT-1. We found that the best-fitting time-lag between swab-positivity and hospital admissions was 19 (18, 20) days and it was 25 (25, 26) days for COVID-19 deaths. The best-fitting population adjusted scaling parameter, a measure of the percentage of people swab-positive who will be in hospital or die after the estimated time lag was 0.24 (0.23, 0.25) for hospitalisations and 0.060 (0.058, 0.062) for deaths. Fewer hospitalisations and COVID-19 deaths occurred from February 2021 after roll-out of the vaccination programme compared to those expected based on the REACT-1 prevalence data, except for hospitalisations during June and July 2021 which coincided with the period during which the Delta variant outcompeted and replaced other strains.

**Figure 3.**
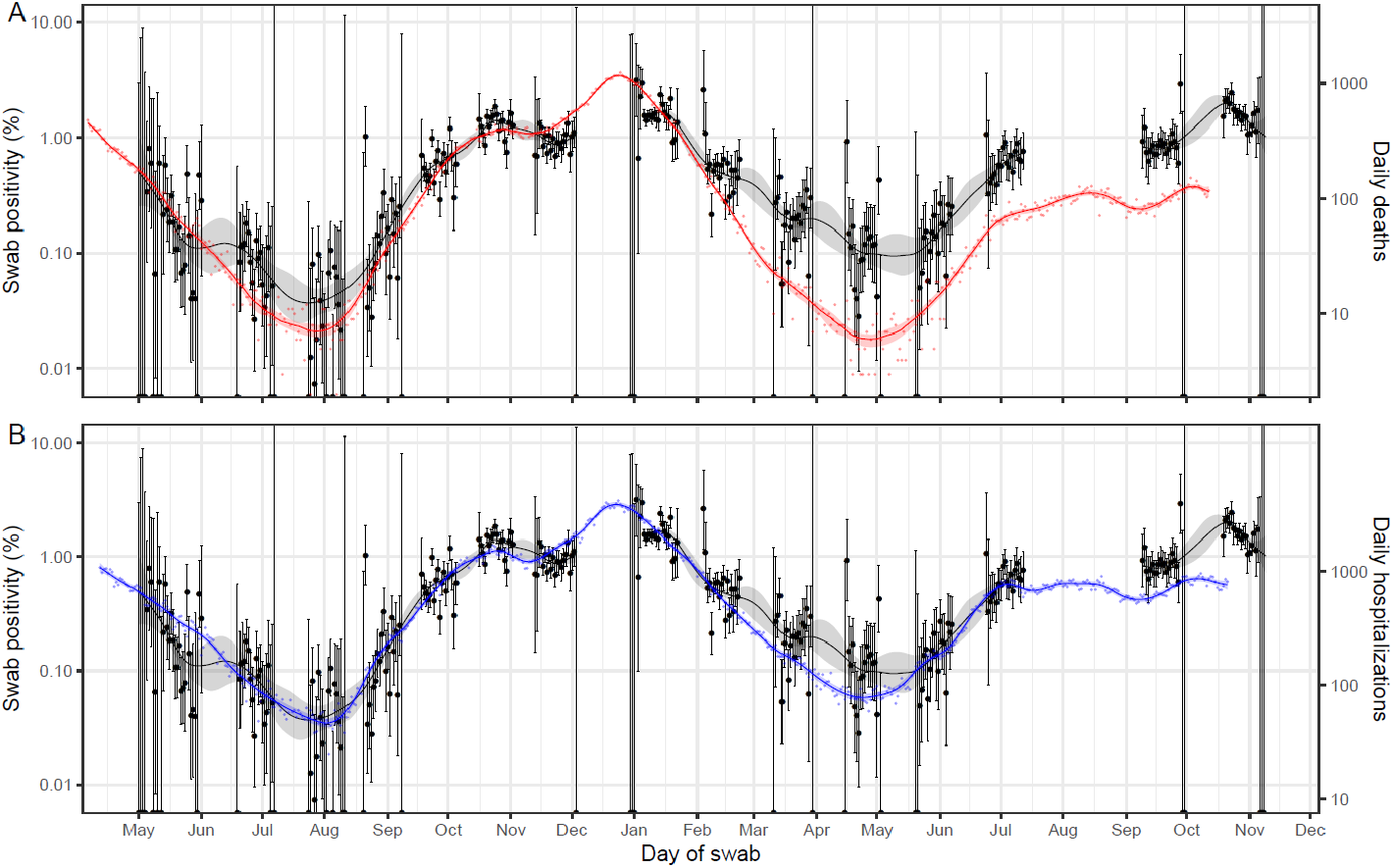
**(A)** comparison of daily deaths and hospitalisations to swab-positivity as measured by REACT-1. Daily swab-positivity for all 15 rounds of the REACT-1 study (black points with 95% confidence intervals, left-hand y-axis) with P-spline estimates for swab-positivity (solid black line, shaded area is 95% credible interval). Daily deaths in England (red points, right-hand y-axis) and P-spline model estimates for expected daily deaths in England (solid red line, shaded area is 95% credible interval, right-hand y-axis). Daily deaths have been shifted by 25 (25, 26) days backwards in time along the x-axis. The daily deaths (right-hand) y-axis has been scaled using the best-fit population adjusted scaling parameter 0.060 (0.058, 0.062). **(B)** Daily hospitalisations in England (blue points, right-hand y-axis) and P-spline model estimates for expected daily hospitalisations in England (solid blue line, shaded area is 95% credible interval, right-hand y-axis). Daily hospitalisations have been shifted by 19 (18, 20) days backwards in time along the x-axis. The daily hospitalisations (right-hand) y-axis has been scaled using the best-fit population adjusted scaling parameter 0.238 (0.230, 0.246).

### Viral sequencing

Sequencing of the positive samples yielded 841 lineages, which were all Delta or Delta sub-lineage variants (Supplementary Table 3). Of these, 57.6% (54.2%, 60.8%, N=484) were AY.4, with AY.4.2 representing 11.8% (9.8%, 14.1%, N=99) of all lineages. This represents a daily growth rate in the proportion of AY.4.2 of 2.80% (1.6%, 4.1%) from round 14 onwards. Compared to AY.4, sub-lineage AY.4.2 was less likely to be associated with symptoms (P=0.04) (Supplementary Table 4-A). Mean Ct values for sub-lineages AY.4 and AY.4.2 were similar suggesting similar mean viral loads (Supplementary Table 4-B)

## Discussion

The REACT-1 programme has been providing timely data on the spread of SARS-CoV-2 in the population of England since lockdown during the first wave of the epidemic in May 2020 [2]. This latest report covers the period from mid-October to early November 2021 spanning the autumn half-term break in schools in England. Compared with the previous round of data collection in September 2021 [11], the prevalence of swab positivity in the population had risen markedly, especially among school-aged children, where we observed rates averaging around 5% over the period. Although prevalence was somewhat lower in older people (65 years and over) it had approximately doubled between rounds, and unlike in younger people, did not appear to be falling during the period of the current study, despite high levels of vaccination in this group.

Our study confirmed that Delta variant and its sub-lineages is the dominant strain circulating in England, as has also been reported by the UK Health Security Agency (UK HSA) based on the national routine testing programme [22]. More than half of the sub-lineages identified corresponded to AY.4, and we found that AY.4.2. was less likely to cause a symptomatic infection. There are ongoing efforts to monitor the spread of that sub-lineage [23,24]. Our data suggest that against the backdrop of Delta and the removal of all restrictions in England, viral transmission increased from September to mid-October 2021, with the subsequent fall in swab positivity being driven by the younger ages, possibly related, at least in part, to the half-term break. The increase in prevalence in older people suggests that transmission can be sustained among the vaccinated as well as the unvaccinated population. A recent study of transmission in the home related to Delta virus suggested only a modest reduction in risk of infection among double-vaccinated compared to unvaccinated individuals, with secondary attack rates of 25% (95% CI 18%, 33%) and 38% (24%, 53%) respectively [5]. In our own data, we found that people living in larger households and those with children in the household had higher prevalence of swab-positivity than single person households and those without children.

In previous rounds we estimated vaccine effectiveness among adults at ages 18 to 64 years [11], but not older adults, because of concern -- given so few were unvaccinated -- that the unvaccinated group might be non-representative of the wider population. Estimating vaccine effectiveness from observational data is well known to run the risk of bias arising from the non-comparability of the vaccinated and unvaccinated groups in key aspects other than vaccination. For example, “confounding by indication” can arise if the presence of one or more underlying conditions affects an individual’s decision to get vaccinated, while a “healthy vaccine bias” may occur where individuals who are healthier are more likely to accede to the vaccination programme [25]. These differential effects can bias estimates of effectiveness in either direction even with statistical adjustments for measured covariates, in an attempt to correct for non-vaccination-related differences between the vaccinated and unvaccinated groups [26]. New approaches are being developed to try to tease out such effects [27].

Our concern about estimating vaccine effectiveness among adults at ages 65 years and over due to non-comparability between the vaccinated and (small) unvaccinated group also became a key consideration here at ages 18 to 64 years. First, reflecting the success of the vaccine programme, only a small proportion of this age group remained unvaccinated in the present round, resulting in wide confidence intervals and imprecise estimates of prevalence in the unvaccinated (comparator) group, especially in the latter rounds. Second, there was possible waning in immunity following vaccination that may have led to increasing risk of breakthrough infections among the double-vaccinated group. Finally, there was evidence that the unvaccinated and double-vaccinated groups differed in important ways according to a variety of parameters, suggesting that the unvaccinated and vaccinated groups were poorly matched.

In contrast, estimates of the effects of the third vaccine dose on swab positivity, compared to two doses, were not affected by these issues in the same way. Since those receiving a third dose of vaccine were already double-vaccinated, they were as a group likely to be much more closely matched to the double-vaccinated group than comparisons of vaccinated and unvaccinated individuals. This was even more so given that the roll-out and scale-up of the booster vaccine programme during the present round meant that many people who wanted to get a third vaccine dose were still waiting their turn. We estimated that following a third vaccine dose, the odds of swab-positivity were on average around one third of the odds of double-vaccinated individuals, indicating an effective immune response from the third dose. However, our estimate is less strong than that reported for symptomatic individuals in both Israel [9] and the UK [28] both of which depended on people presenting with symptoms to the national testing programmes -- whereas our estimate is community-based, includes non-symptomatic individuals and is not contingent on test-seeking behaviours.

The vaccinated and unvaccinated groups of children aged 12 to 17 were also likely to be much more comparable to each other in round 15 (19 October to 5 November 2021) than were the vaccinated and unvaccinated groups of adults, because many schools had yet to be visited by vaccination teams so many unvaccinated children had not yet had the opportunity to be vaccinated. Our results clearly show the benefits of single doses delivered to children aged 12 years and above in terms of reducing the risk of swab positivity and by extension onward transmission of infection. Preventing infections is important because, as already noted above, those who have breakthrough infections post-vaccination do transmit to others, including within households at a rate comparable to those who are infected but have not been vaccinated [5]. In addition, high rates of infections in schools are disruptive to learning and education.

Crucially, the vaccination programme has extensively reduced the risks of COVID-19 related hospitalisations and deaths [29]. In January 2021 when the average swab-positivity prevalence in REACT-1 was similar to what was recorded in the current round, hospitalisations in England were running at around 3,500 per day compared with around 850 per day at the beginning of November 2021 [30]. Looking further at our own data, we can observe a weakening of the association between swab-positivity in REACT-1 and COVID-19 related hospitalisations and deaths arising on average 19 and 25 days later. Nonetheless it remains essential that the booster programme in the UK is rapidly able to reach the vast majority of the more vulnerable population to prevent further pressure on health services from waning immunity.

Our surveys have limitations. Although the response rate in REACT-1 has declined steadily from 30.5% in round 1 to its current level of 11.7% in round 15, we use rim weighting to obtain prevalence estimates that are representative of the population of England as a whole. While we obtain vaccination data reported by participants, here we relied on data on vaccination as recorded by the NHS for the ∼87% (round 15) who gave permission to link to their NHS records. This has the advantage of providing accurate information on the date of vaccination and type of vaccine used, both important to avoid errors and biases in estimates of prevalence and vaccine effectiveness, and there is no dependency on participant recall. However, to the extent that those who consent and do not consent to data linkage may differ, it is possible that undetected systematic errors may be introduced (this possibility is lessened by the large proportion of people who do consent to linkage). Finally, we changed the method by which swabs were sent to the laboratory for RT-PCR in the present round, relying on the priority postal service for participants to return their swab (transported in saline solution). In the previous round we tested in a 1:1 randomised fashion either courier pick-up (no cold chain) or priority postal service and found no discernible difference between the two in either positivity rate or Ct values [11]. Prior to that, dry swabs were sent by courier to the laboratory on a cold chain.

In conclusion, swab-positivity was very high at the start of round 15, reaching a maximum around 20 to 21 October 2021, and then falling through late October with an uncertain trend in the last few days of data collection. School-aged children were the most likely to test positive, while at the same time those children who had received a single dose of the Pfizer-BioNTech vaccine at least 14 days prior to swabbing were at around 50% risk of infection compared to unvaccinated students of the same age (12 to 17 years of age). Likewise, booster (third) vaccine doses were found to protect adults from infection, compared to their counterparts who had received only two doses. The relatively small proportion of adults who remain unvaccinated (a clear positive from a public health perspective) limits our ability to reliably estimate vaccine effectiveness comparing those who have received two doses of the vaccine to those remaining unvaccinated. Thus, we cannot usefully add to the findings from round 14 [11] that showed the protection offered by two vaccine doses waned over time. Expanded availability and rapid roll-out of booster doses, second doses for teenagers aged 16 to 17 years and single doses for children aged 12 to 15 years in England, with possible extension to younger children, should help reduce transmission during the winter period when healthcare demands typically rise.

## Data Availability

Access to REACT-1 data is restricted due to ethical and security considerations.
Summary statistics and descriptive tables from the current REACT-1 study are available in the Supplementary Information.
Additional summary statistics and results from the REACT-1 programme are also available at
https://www.imperial.ac.uk/medicine/research-and-impact/groups/react-study/real-time-assessment-of-community-transmission-findings/
and
https://github.com/mrc-ide/reactidd/tree/master/inst/extdata REACT-1
Study Materials are available for each round at
https://www.imperial.ac.uk/medicine/research-and-impact/groups/react-study/react-1-study-materials/
Sequence read data are available without restriction from the European Nucleotide Archive at
https://www.ebi.ac.uk/ena/browser/view/PRJEB37886, and consensus genome sequences are available from the Global initiative on sharing all influenza data at
https://www.gisaid.org

## Ethics

We obtained research ethics approval from the South Central-Berkshire B Research Ethics Committee (IRAS ID: 283787).

## Contributors

PE and CAD are corresponding authors. PE, MC-H and CAD conceived the study and the analytical plan. MC-H, OE, BB, HWang, DH, JJ and CEW performed the statistical analyses. HWang, OE, DH, BB, and MW curated the data. CA, PJD, DA, WB, GT, GC, HW, AD provided study oversight and results interpretation. AJP generated the sequencing data. AD and PE obtained funding. All authors revised the manuscript for important intellectual content and approved the submission of the manuscript. PE had full access to the data and takes responsibility for the integrity of the data and the accuracy of the data analysis, and for the decision to submit for publication.

## Acknowledgements

MC-H and MW acknowledge support from the H2020-EXPANSE project (Horizon 2020 grant No 874627). MC-H and BB acknowledge support from Cancer Research UK, Population Research Committee Project grant ‘Mechanomics’ (grant No 22184 to MC-H). CAD acknowledges support from the MRC Centre for Global Infectious Disease Analysis and National Institute for Health Research (NIHR) Health Protection Research Unit (HPRU). GC is supported by an NIHR Professorship. HW acknowledges support from an NIHR Senior Investigator Award and the Wellcome Trust (205456/Z/16/Z). PE is Director of the Medical Research Council (MRC) Centre for Environment and Health (MR/L01341X/1, MR/S019669/1). PE acknowledges support from Health Data Research UK (HDR UK); the NIHR Imperial Biomedical Research Centre; NIHR Health Protection Research Units in Chemical and Radiation Threats and Hazards, and Environmental Exposures and Health; the British Heart Foundation Centre for Research Excellence at Imperial College London (RE/18/4/34215); and the UK Dementia Research Institute at Imperial College London (MC_PC_17114). AJP acknowledges the support of the Biotechnology and Biological Sciences Research Council (BB/R012504/1). We thank The Huo Family Foundation for their support of our work on COVID-19.

We thank key collaborators on this work – Ipsos MORI: Kelly Beaver, Sam Clemens, Gary Welch, Nicholas Gilby, Kelly Ward, Galini Pantelidou and Kevin Pickering; Institute of Global Health Innovation at Imperial College London: Gianluca Fontana, Justine Alford; School of Public Health, Imperial College London: Eric Johnson, Rob Elliott, Graham Blakoe; Quadram Institute, Norwich, UK: Alexander J. Trotter; North West London Pathology and Public Health England (now UKHSA) for help in calibration of the laboratory analyses; Patient Experience Research Centre at Imperial College London and the REACT Public Advisory Panel; NHS Digital for access to the NHS register; the Department of Health and Social Care for logistic support.

## Data Sharing

Access to REACT-1 data is restricted due to ethical and security considerations. Summary statistics and descriptive tables from the current REACT-1 study are available in the Supplementary Information. Additional summary statistics and results from the REACT-1 programme are also available at https://www.imperial.ac.uk/medicine/research-and-impact/groups/react-study/real-time-assessment-of-community-transmission-findings/ and https://github.com/mrc-ide/reactidd/tree/master/inst/extdata REACT-1 Study Materials are available for each round at https://www.imperial.ac.uk/medicine/research-and-impact/groups/react-study/react-1-study-materials/

Sequence read data are available without restriction from the European Nucleotide Archive at https://www.ebi.ac.uk/ena/browser/view/PRJEB37886, and consensus genome sequences are available from the Global initiative on sharing all influenza data at https://www.gisaid.org.

## Supplementary Tables and Figures

**Supplementary Table 1.** Unweighted and weighted prevalence of swab-positivity from REACT-1 across rounds 1 to 15.

| Round | Tested swabs | Positive swabs | Unweighted prevalence (95% CI) | Weighted prevalence (95% CI) | First sample | Last sample |
| --- | --- | --- | --- | --- | --- | --- |
| 1 | 120,620 | 159 | 0.13% (0.11%, 0.15%) | 0.16% (0.13%, 0.19%) | 01/05/20 | 01/06/20 |
| 2 | 159,199 | 123 | 0.08% (0.07%, 0.09%) | 0.09% (0.07%, 0.11%) | 19/06/20 | 07/07/20 |
| 3 | 162,821 | 54 | 0.03% (0.03%, 0.04%) | 0.04% (0.03%, 0.05%) | 24/07/20 | 11/08/20 |
| 4 | 154,325 | 137 | 0.09% (0.08%, 0.11%) | 0.13% (0.01%, 0.15%) | 20/08/20 | 08/09/20 |
| 5 | 174,949 | 824 | 0.47% (0.44%, 0.50%) | 0.60% (0.55%, 0.71%) | 18/09/20 | 05/10/20 |
| 6 | 160,175 | 1,732 | 1.08% (1.03%, 1.13%) | 1.30% (1.21%, 1.39%) | 16/10/20 | 02/11/20 |
| 7 | 168,181 | 1,299 | 0.77% (0.73%, 0.82%) | 0.94% (0.87%, 1.01%) | 13/11/20 | 03/12/20 |
| 8 | 167,642 | 2,282 | 1.36% (1.31%, 1.42%) | 1.57% (1.49%, 1.66%) | 06/01/21 | 22/01/21 |
| 9 | 165,456 | 689 | 0.42% (0.39%, 0.45%) | 0.49% (0.44%, 0.55%) | 04/02/21 | 23/02/21 |
| 10 | 140,844 | 227 | 0.16% (0.14%, 0.18%) | 0.20% (0.17%, 0.23%) | 11/03/21 | 30/03/21 |
| 11 | 127,408 | 115 | 0.09% (0.07%, 0.11%) | 0.10% (0.08%, 0.13%) | 15/04/21 | 03/05/21 |
| 12* | 108,911 | 135 | 0.12% (0.10%, 0.15%) | 0.15% (0.12%, 0.18%) | 20/05/21 | 07/06/21 |
| 13 | 98,233 | 527 | 0.54% (0.49%, 0.58%) | 0.63% (0.57%, 0.69%) | 24/06/21 | 12/07/21 |
| 14** | 100,527 | 764 | 0.76% (0.71%, 0.82%) | 0.83% (0.76%, 0.89%) | 09/09/21 | 27/09/21 |
| 15*** | 100,112 | 1,399 | 1.40% (1.33%, 1.47%) | 1.57% (1.48%, 1.66%) | 19/10/21 | 05/11/21 |

\* Sampling strategy changed for round 12 and subsequent rounds. Therefore unweighted prevalence is not directly comparable with previous rounds
\*\* Including N=509 samples from 28-30 September. Sample handling changed in round 14. Prevalence is not directly comparable with previous rounds
\*\*\* Including N=93 samples (all negatives) from 6-8 November, and N=86 samples with no collection/arrival dates

**Supplementary Table 2a.**
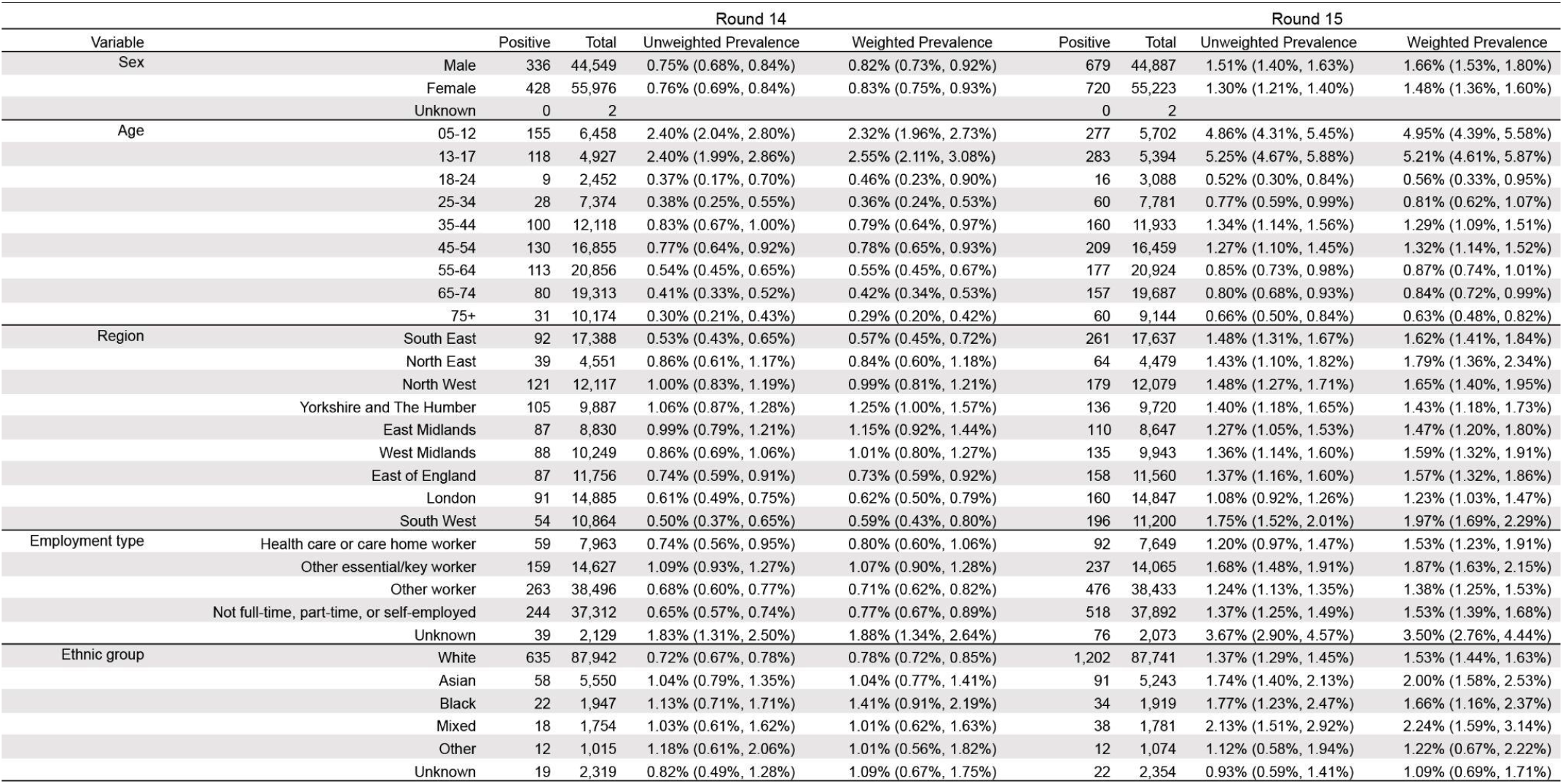
Unweighted and weighted prevalence of swab-positivity in round 14 and round 15 by sex, age, region, employment type, and ethnic group.

| Variable |  | Round 14 |  |  |  | Round 15 |  |  |  |
| --- | --- | --- | --- | --- | --- | --- | --- | --- | --- |
|  |  | Positive | Total | Unweighted Prevalence | Weighted Prevalence | Positive | Total | Unweighted Prevalence | Weighted Prevalence |
| Sex | Male | 336 | 44,549 | 0.75% (0.68%, 0.84%) | 0.82% (0.73%, 0.92%) | 679 | 44,887 | 1.51% (1.40%, 1.63%) | 1.66% (1.53%, 1.80%) |
|  | Female | 428 | 55,976 | 0.76% (0.69%, 0.84%) | 0.83% (0.75%, 0.93%) | 720 | 55,223 | 1.30% (1.21%, 1.40%) | 1.48% (1.36%, 1.60%) |
|  | Unknown | 0 | 2 |  |  | 0 | 2 |  |  |
| Age | 05-12 | 155 | 6,458 | 2.40% (2.04%, 2.80%) | 2.32% (1.96%, 2.73%) | 277 | 5,702 | 4.86% (4.31%, 5.45%) | 4.95% (4.39%, 5.58%) |
|  | 13-17 | 118 | 4,927 | 2.40% (1.99%, 2.86%) | 2.55% (2.11%, 3.08%) | 283 | 5,394 | 5.25% (4.67%, 5.88%) | 5.21% (4.61%, 5.87%) |
|  | 18-24 | 9 | 2,452 | 0.37% (0.17%, 0.70%) | 0.46% (0.23%, 0.90%) | 16 | 3,088 | 0.52% (0.30%, 0.84%) | 0.56% (0.33%, 0.95%) |
|  | 25-34 | 28 | 7,374 | 0.38% (0.25%, 0.55%) | 0.36% (0.24%, 0.53%) | 60 | 7,781 | 0.77% (0.59%, 0.99%) | 0.81% (0.62%, 1.07%) |
|  | 35-44 | 100 | 12,118 | 0.83% (0.67%, 1.00%) | 0.79% (0.64%, 0.97%) | 160 | 11,933 | 1.34% (1.14%, 1.56%) | 1.29% (1.09%, 1.51%) |
|  | 45-54 | 130 | 16,855 | 0.77% (0.64%, 0.92%) | 0.78% (0.65%, 0.93%) | 209 | 16,459 | 1.27% (1.10%, 1.45%) | 1.32% (1.14%, 1.52%) |
|  | 55-64 | 113 | 20,856 | 0.54% (0.45%, 0.65%) | 0.55% (0.45%, 0.67%) | 177 | 20,924 | 0.85% (0.73%, 0.98%) | 0.87% (0.74%, 1.01%) |
|  | 65-74 | 80 | 19,313 | 0.41% (0.33%, 0.52%) | 0.42% (0.34%, 0.53%) | 157 | 19,687 | 0.80% (0.68%, 0.93%) | 0.84% (0.72%, 0.99%) |
|  | 75+ | 31 | 10,174 | 0.30% (0.21%, 0.43%) | 0.29% (0.20%, 0.42%) | 60 | 9,144 | 0.66% (0.50%, 0.84%) | 0.63% (0.48%, 0.82%) |
| Region | South East | 92 | 17,388 | 0.53% (0.43%, 0.65%) | 0.57% (0.45%, 0.72%) | 261 | 17,637 | 1.48% (1.31%, 1.67%) | 1.62% (1.41%, 1.84%) |
|  | North East | 39 | 4,551 | 0.86% (0.61%, 1.17%) | 0.84% (0.60%, 1.18%) | 64 | 4,479 | 1.43% (1.10%, 1.82%) | 1.79% (1.36%, 2.34%) |
|  | North West | 121 | 12,117 | 1.00% (0.83%, 1.19%) | 0.99% (0.81%, 1.21%) | 179 | 12,079 | 1.48% (1.27%, 1.71%) | 1.65% (1.40%, 1.95%) |
|  | Yorkshire and The Humber | 105 | 9,887 | 1.06% (0.87%, 1.28%) | 1.25% (1.00%, 1.57%) | 136 | 9,720 | 1.40% (1.18%, 1.65%) | 1.43% (1.18%, 1.73%) |
|  | East Midlands | 87 | 8,830 | 0.99% (0.79%, 1.21%) | 1.15% (0.92%, 1.44%) | 110 | 8,647 | 1.27% (1.05%, 1.53%) | 1.47% (1.20%, 1.80%) |
|  | West Midlands | 88 | 10,249 | 0.86% (0.69%, 1.06%) | 1.01% (0.80%, 1.27%) | 135 | 9,943 | 1.36% (1.14%, 1.60%) | 1.59% (1.32%, 1.91%) |
|  | East of England | 87 | 11,756 | 0.74% (0.59%, 0.91%) | 0.73% (0.59%, 0.92%) | 158 | 11,560 | 1.37% (1.16%, 1.60%) | 1.57% (1.32%, 1.86%) |
|  | London | 91 | 14,885 | 0.61% (0.49%, 0.75%) | 0.62% (0.50%, 0.79%) | 160 | 14,847 | 1.08% (0.92%, 1.26%) | 1.23% (1.03%, 1.47%) |
|  | South West | 54 | 10,864 | 0.50% (0.37%, 0.65%) | 0.59% (0.43%, 0.80%) | 196 | 11,200 | 1.75% (1.52%, 2.01%) | 1.97% (1.69%, 2.29%) |
| Employment type | Health care or care home worker | 59 | 7,963 | 0.74% (0.56%, 0.95%) | 0.80% (0.60%, 1.06%) | 92 | 7,649 | 1.20% (0.97%, 1.47%) | 1.53% (1.23%, 1.91%) |
|  | Other essential/key worker | 159 | 14,627 | 1.09% (0.93%, 1.27%) | 1.07% (0.90%, 1.28%) | 237 | 14,065 | 1.68% (1.48%, 1.91%) | 1.87% (1.63%, 2.15%) |
|  | Other worker | 263 | 38,496 | 0.68% (0.60%, 0.77%) | 0.71% (0.62%, 0.82%) | 476 | 38,433 | 1.24% (1.13%, 1.35%) | 1.38% (1.25%, 1.53%) |
|  | Not full-time, part-time, or self-employed | 244 | 37,312 | 0.65% (0.57%, 0.74%) | 0.77% (0.67%, 0.89%) | 518 | 37,892 | 1.37% (1.25%, 1.49%) | 1.53% (1.39%, 1.68%) |
|  | Unknown | 39 | 2,129 | 1.83% (1.31%, 2.50%) | 1.88% (1.34%, 2.64%) | 76 | 2,073 | 3.67% (2.90%, 4.57%) | 3.50% (2.76%, 4.44%) |
| Ethnic group | White | 635 | 87,942 | 0.72% (0.67%, 0.78%) | 0.78% (0.72%, 0.85%) | 1,202 | 87,741 | 1.37% (1.29%, 1.45%) | 1.53% (1.44%, 1.63%) |
|  | Asian | 58 | 5,550 | 1.04% (0.79%, 1.35%) | 1.04% (0.77%, 1.41%) | 91 | 5,243 | 1.74% (1.40%, 2.13%) | 2.00% (1.58%, 2.53%) |
|  | Black | 22 | 1,947 | 1.13% (0.71%, 1.71%) | 1.41% (0.91%, 2.19%) | 34 | 1,919 | 1.77% (1.23%, 2.47%) | 1.66% (1.16%, 2.37%) |
|  | Mixed | 18 | 1,754 | 1.03% (0.61%, 1.62%) | 1.01% (0.62%, 1.63%) | 38 | 1,781 | 2.13% (1.51%, 2.92%) | 2.24% (1.59%, 3.14%) |
|  | Other | 12 | 1,015 | 1.18% (0.61%, 2.06%) | 1.01% (0.56%, 1.82%) | 12 | 1,074 | 1.12% (0.58%, 1.94%) | 1.22% (0.67%, 2.22%) |
|  | Unknown | 19 | 2,319 | 0.82% (0.49%, 1.28%) | 1.09% (0.67%, 1.75%) | 22 | 2,354 | 0.93% (0.59%, 1.41%) | 1.09% (0.69%, 1.71%) |

**Supplementary Table 2b.** Unweighted and weighted prevalence of swab-positivity in round 14 and round 15 by household size, number of children in the household, contact with a COVID-19 case, symptom status and neighbourhood deprivation.

|  |  | Round 14 |  |  |  | Round 15 |  |  |  |
| --- | --- | --- | --- | --- | --- | --- | --- | --- | --- |
| Variable |  | Positive | Total | Unweighted Prevalence | Weighted Prevalence | Positive | Total | Unweighted Prevalence | Weighted Prevalence |
| Household size | 1 | 57 | 16,613 | 0.34% (0.26%, 0.44%) | 0.33% (0.25%, 0.44%) | 132 | 16,997 | 0.78% (0.65%, 0.92%) | 0.78% (0.65%, 0.94%) |
|  | 2 | 174 | 39,044 | 0.45% (0.38%, 0.52%) | 0.46% (0.39%, 0.54%) | 318 | 39,624 | 0.80% (0.72%, 0.90%) | 0.83% (0.74%, 0.94%) |
|  | 3 | 150 | 17,235 | 0.87% (0.74%, 1.02%) | 0.93% (0.78%, 1.10%) | 283 | 17,579 | 1.61% (1.43%, 1.81%) | 1.76% (1.54%, 2.00%) |
|  | 4 | 250 | 19,154 | 1.31% (1.15%, 1.48%) | 1.32% (1.15%, 1.51%) | 421 | 17,774 | 2.37% (2.15%, 2.60%) | 2.48% (2.23%, 2.74%) |
|  | 5 | 89 | 6,057 | 1.47% (1.18%, 1.81%) | 1.41% (1.12%, 1.76%) | 178 | 5,752 | 3.09% (2.66%, 3.58%) | 3.12% (2.66%, 3.66%) |
|  | 6+ | 44 | 2,424 | 1.82% (1.32%, 2.43%) | 1.75% (1.24%, 2.46%) | 67 | 2,386 | 2.81% (2.18%, 3.55%) | 2.95% (2.27%, 3.82%) |
| Number of children in the household | 0 | 276 | 66,025 | 0.42% (0.37%, 0.47%) | 0.40% (0.35%, 0.46%) | 491 | 66,590 | 0.74% (0.67%, 0.81%) | 0.74% (0.67%, 0.81%) |
|  | 1+ | 378 | 28,659 | 1.32% (1.19%, 1.46%) | 1.37% (1.22%, 1.52%) | 644 | 27,239 | 2.36% (2.19%, 2.55%) | 2.62% (2.41%, 2.85%) |
|  | Unknown | 110 | 5,843 | 1.88% (1.55%, 2.26%) | 2.06% (1.69%, 2.51%) | 264 | 6,283 | 4.20% (3.72%, 4.73%) | 4.22% (3.72%, 4.78%) |
| COVID case contact | No | 325 | 80,587 | 0.40% (0.36%, 0.45%) | 0.43% (0.38%, 0.49%) | 595 | 80,225 | 0.74% (0.68%, 0.80%) | 0.83% (0.75%, 0.90%) |
|  | Yes, contact with a confirmed/tested COVID-19 case | 293 | 4,044 | 7.25% (6.47%, 8.09%) | 7.35% (6.50%, 8.31%) | 554 | 6,509 | 8.51% (7.84%, 9.22%) | 9.13% (8.35%, 9.96%) |
|  | Yes, contact with a suspected COVID-19 case | 38 | 1,058 | 3.59% (2.55%, 4.90%) | 3.80% (2.70%, 5.32%) | 67 | 1,448 | 4.63% (3.60%, 5.84%) | 5.04% (3.88%, 6.51%) |
|  | Unknown | 108 | 14,838 | 0.73% (0.60%, 0.88%) | 0.79% (0.65%, 0.97%) | 183 | 11,930 | 1.53% (1.32%, 1.77%) | 1.63% (1.39%, 1.91%) |
| Symptom status | Classic COVID symptoms* | 359 | 4,963 | 7.23% (6.53%, 7.99%) | 6.85% (6.12%, 7.68%) | 655 | 8,351 | 7.84% (7.28%, 8.44%) | 7.84% (7.22%, 8.51%) |
|  | Other symptoms | 108 | 11,337 | 0.95% (0.78%, 1.15%) | 0.95% (0.77%, 1.17%) | 208 | 15,634 | 1.33% (1.16%, 1.52%) | 1.47% (1.27%, 1.70%) |
|  | No symptoms | 190 | 69,446 | 0.27% (0.24%, 0.32%) | 0.31% (0.27%, 0.37%) | 357 | 64,231 | 0.56% (0.50%, 0.62%) | 0.67% (0.60%, 0.75%) |
|  | Unknown | 107 | 14,781 | 0.72% (0.59%, 0.87%) | 0.79% (0.64%, 0.97%) | 179 | 11,896 | 1.50% (1.29%, 1.74%) | 1.60% (1.36%, 1.88%) |
| Deprivation | 1 Most deprived | 119 | 11,777 | 1.01% (0.84%, 1.21%) | 0.98% (0.81%, 1.20%) | 174 | 11,556 | 1.51% (1.29%, 1.74%) | 1.70% (1.45%, 1.99%) |
|  | 2 | 118 | 16,962 | 0.70% (0.58%, 0.83%) | 0.75% (0.61%, 0.92%) | 215 | 16,850 | 1.28% (1.11%, 1.46%) | 1.41% (1.22%, 1.63%) |
|  | 3 | 142 | 21,117 | 0.67% (0.57%, 0.79%) | 0.75% (0.63%, 0.90%) | 283 | 21,096 | 1.34% (1.19%, 1.51%) | 1.51% (1.33%, 1.72%) |
|  | 4 | 178 | 24,102 | 0.74% (0.63%, 0.85%) | 0.76% (0.65%, 0.89%) | 327 | 24,130 | 1.36% (1.21%, 1.51%) | 1.55% (1.38%, 1.74%) |
|  | 5 Least deprived | 207 | 26,569 | 0.78% (0.68%, 0.89%) | 0.90% (0.77%, 1.04%) | 400 | 26,480 | 1.51% (1.37%, 1.66%) | 1.67% (1.51%, 1.86%) |

\* Classic COVID symptoms: loss or change of sense of smell or taste, fever, new persistent cough

**Supplementary Table 3.** Proportion of each Delta sub-lineage detected in 841 positive samples from round 15.

| Sub-lineage | N (841) | Proportion |
| --- | --- | --- |
| B.1.617.2 (Delta/Sub-lineage not known) | 108 | 0.128 ( 0.107 , 0.153 ) |
| AY.109 | 1 | 0.001 ( 0.000 , 0.007 ) |
| AY.110 | 1 | 0.001 ( 0.000 , 0.007 ) |
| AY.111 | 3 | 0.004 ( 0.001 , 0.010 ) |
| AY.25 | 1 | 0.001 ( 0.000 , 0.007 ) |
| AY.33 | 1 | 0.001 ( 0.000 , 0.007 ) |
| AY.34 | 6 | 0.007 ( 0.003 , 0.015 ) |
| AY.39 | 7 | 0.008 ( 0.004 , 0.017 ) |
| AY.4 | 484 | 0.576 ( 0.542 , 0.608 ) |
| AY.4.2 | 99 | 0.118 ( 0.098 , 0.141 ) |
| AY.4.5 | 4 | 0.005 ( 0.002 , 0.012 ) |
| AY.42 | 5 | 0.006 ( 0.003 , 0.014 ) |
| AY.43 | 40 | 0.048 ( 0.035 , 0.064 ) |
| AY.44 | 13 | 0.015 ( 0.009 , 0.026 ) |
| AY.46.2 | 3 | 0.004 ( 0.001 , 0.010 ) |
| AY.46.5 | 6 | 0.007 ( 0.003 , 0.015 ) |
| AY.47 | 2 | 0.002 ( 0.001 , 0.009 ) |
| AY.5 | 21 | 0.025 ( 0.016 , 0.038 ) |
| AY.5.4 | 1 | 0.001 ( 0.000 , 0.007 ) |
| AY.57 | 1 | 0.001 ( 0.000 , 0.007 ) |
| AY.6 | 13 | 0.015 ( 0.009 , 0.026 ) |
| AY.60 | 1 | 0.001 ( 0.000 , 0.007 ) |
| AY.7.2 | 1 | 0.001 ( 0.000 , 0.007 ) |
| AY.79 | 1 | 0.001 ( 0.000 , 0.007 ) |
| AY.80 | 1 | 0.001 ( 0.000 , 0.007 ) |
| AY.88 | 1 | 0.001 ( 0.000 , 0.007 ) |
| AY.9 | 2 | 0.002 ( 0.001 , 0.009 ) |
| AY.9.1 | 2 | 0.002 ( 0.001 , 0.009 ) |
| AY.9.2 | 1 | 0.001 ( 0.000 , 0.007 ) |
| AY.90 | 1 | 0.001 ( 0.000 , 0.007 ) |
| AY.91 | 1 | 0.001 ( 0.000 , 0.007 ) |
| AY.98 | 9 | 0.011 ( 0.006 , 0.020 ) |

**Supplementary Table 4-A.** Reported symptoms in swab positive participants by B.1.617.2 and sub-lineages with N=10 or more

| Sub-lineage | N | Any symptoms | Proportion | P-value | Classic COVID-19 symptoms* | Proportion | P-value |
| --- | --- | --- | --- | --- | --- | --- | --- |
| AY.4 | 484 | 370 | 0.764 ( 0.725 , 0.800 ) | ref | 224 | 0.463 ( 0.419 , 0.507 ) | ref |
| AY.4.2 | 99 | 66 | 0.667 ( 0.569 , 0.752 ) | 0.04 | 33 | 0.333 ( 0.248 , 0.431 ) | 0.02 |
| AY.43 | 40 | 34 | 0.850 ( 0.709 , 0.929 ) | 0.22 | 23 | 0.575 ( 0.422 , 0.715 ) | 0.17 |
| AY.44 | 13 | 9 | 0.692 ( 0.424 , 0.873 ) | 0.55 | 7 | 0.538 ( 0.291 , 0.768 ) | 0.59 |
| AY.5 | 21 | 17 | 0.810 ( 0.600 , 0.923 ) | 0.63 | 13 | 0.619 ( 0.409 , 0.792 ) | 0.17 |
| AY.6 | 13 | 11 | 0.846 ( 0.578 , 0.957 ) | 0.50 | 7 | 0.538 ( 0.291 , 0.768 ) | 0.59 |
| B.1.617.2 | 108 | 83 | 0.769 ( 0.681 , 0.838 ) | 0.93 | 50 | 0.463 ( 0.372 , 0.557 ) | 1.00 |

\* Classic COVID symptoms: loss or change of sense of smell or taste, fever, new persistent cough

**Supplementary Table 4-B.** Mean Ct values for N-gene and E-gene by B.1.617.2 and sub-lineages with N=10 or more.

| Sub-lineage | Mean N-gene Ct value | P-value | Mean E-gene Ct value | P-value |
| --- | --- | --- | --- | --- |
| AY.4 | 24.19 ( 23.82 , 24.56 ) | ref | 22.70 ( 22.33 , 23.07 ) | ref |
| B.1.617.2 | 25.64 ( 24.86 , 26.43 ) | <0.01 | 24.44 ( 23.66 , 25.22 ) | <0.01 |
| AY.5 | 24.13 ( 22.36 , 25.90 ) | 0.94 | 22.71 ( 20.93 , 24.48 ) | 0.99 |
| AY.6 | 22.96 ( 20.71 , 25.21 ) | 0.29 | 21.56 ( 19.30 , 23.81 ) | 0.33 |
| AY.4.2 | 23.64 ( 22.82 , 24.45 ) | 0.23 | 22.17 ( 21.35 , 22.98 ) | 0.25 |
| AY.43 | 25.33 ( 24.05 , 26.61 ) | 0.09 | 23.67 ( 22.38 , 24.95 ) | 0.16 |
| AY.44 | 28.60 ( 26.35 , 30.85 ) | <0.01 | 27.70 ( 25.44 , 29.95 ) | <0.01 |

**Supplementary Figure 1-A.**
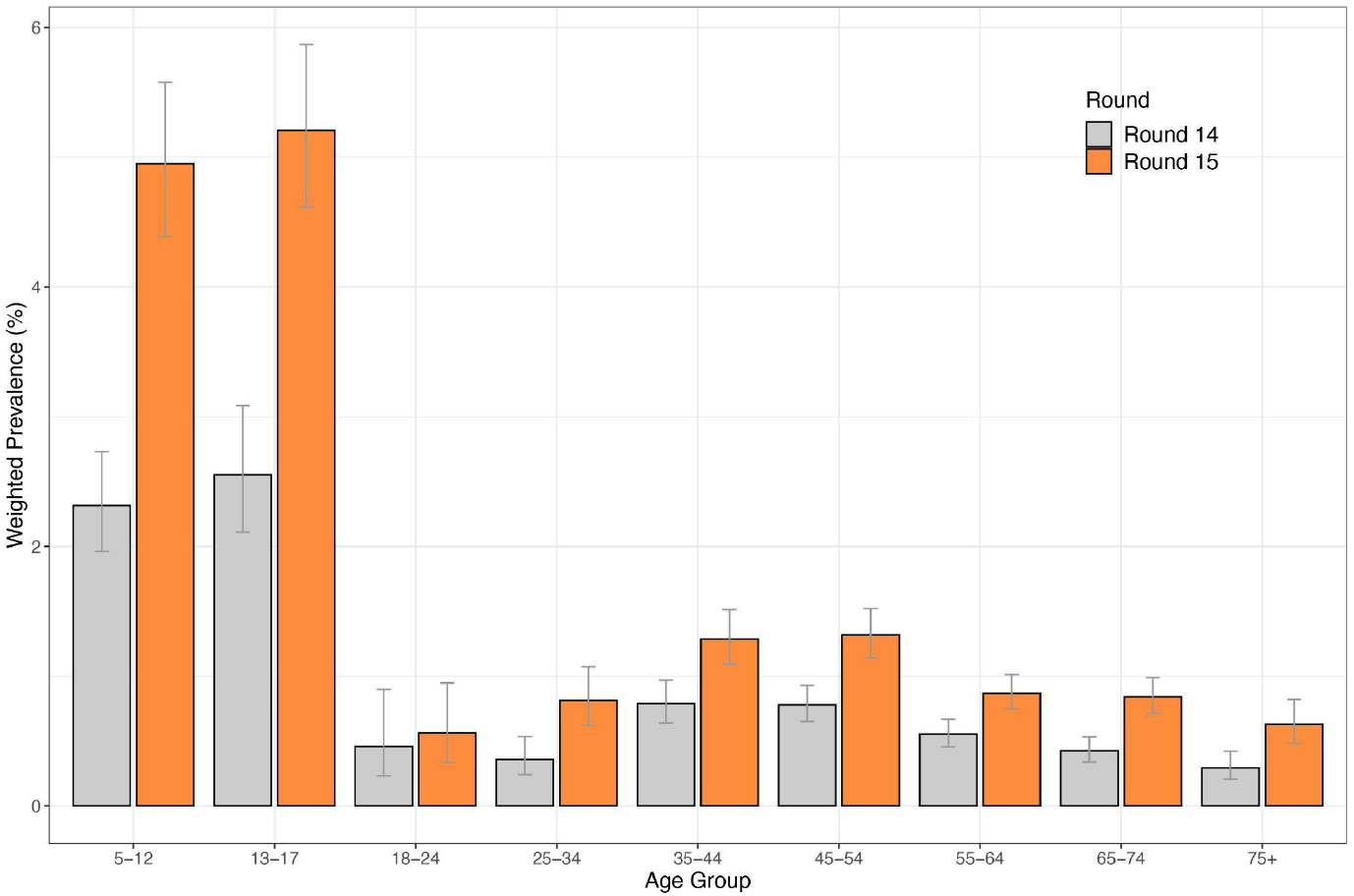
Weighted prevalence of swab-positivity by age group for round 14 and round 15. Bars show the prevalence point estimates (grey for round 14 and orange for round 15), and the vertical lines represent the 95% confidence intervals.

**Supplementary Figure 1-B.**
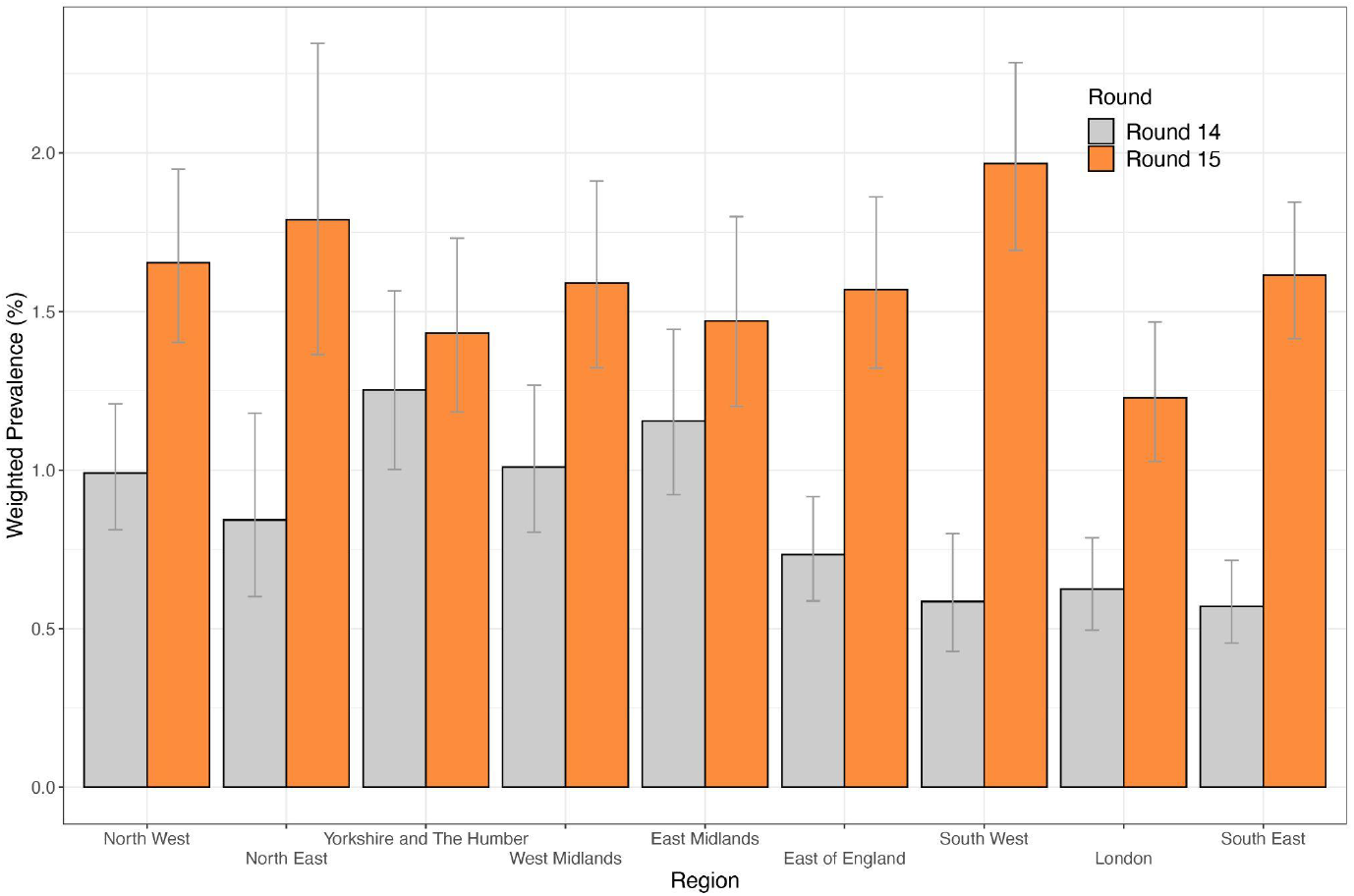
Weighted prevalence of swab-positivity by region for round 14 and round 15. Bars represent prevalence point estimates (grey for round 14 and orange for round 15), and the vertical lines the corresponding 95% confidence intervals.

**Supplementary Figure 2.**
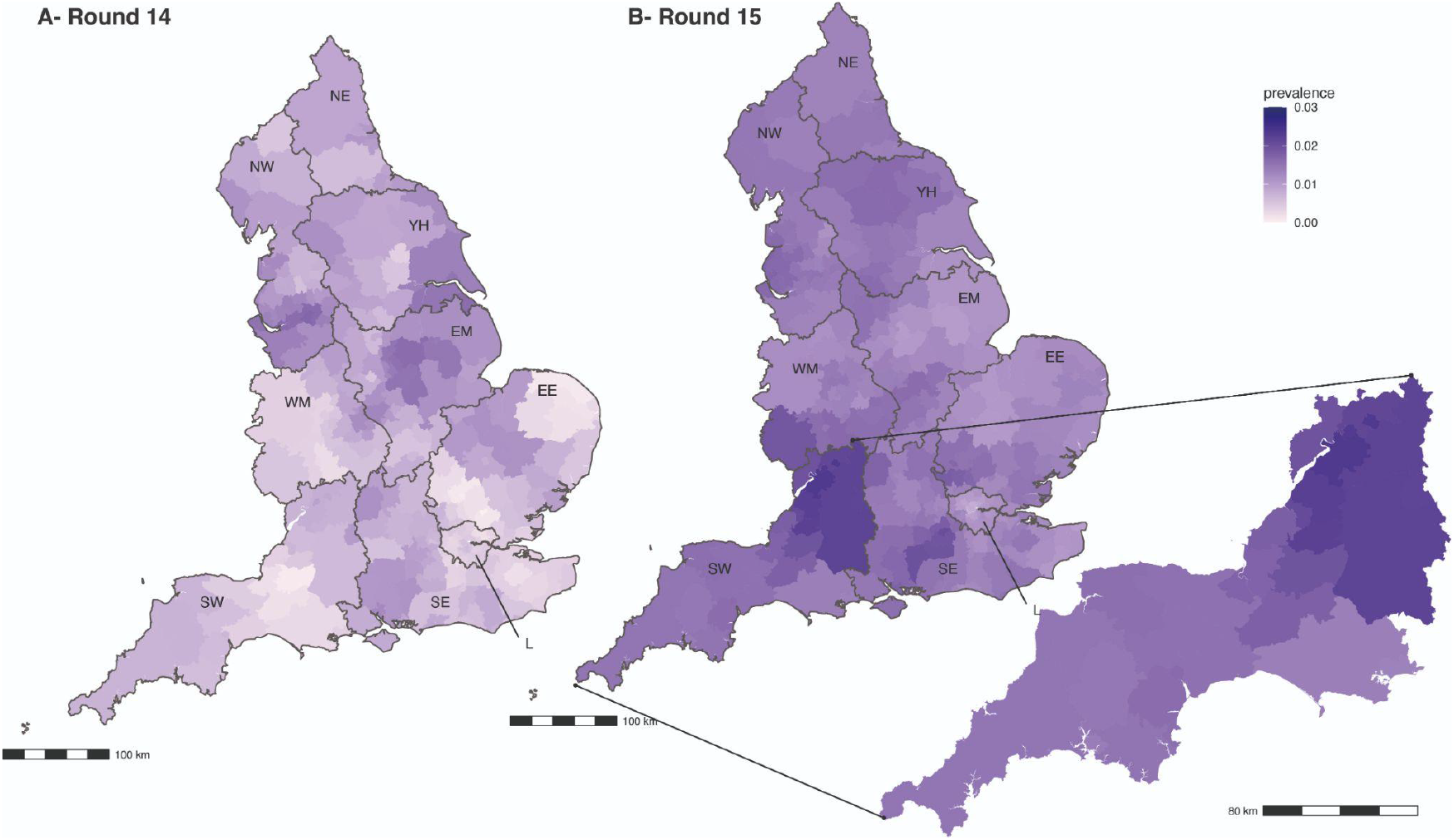
Neighbourhood smoothed average prevalence by lower tier local authority area for (A) round 14 and (B) round 15. Neighbourhood prevalence calculated from nearest neighbours (the median number of neighbours within 30 km in the study). Average neighbourhood prevalence displayed for individual lower-tier local authorities. Regions: NE = North East, NW = North West, YH = Yorkshire and The Humber, EM = East Midlands, WM = West Midlands, EE = East of England, L = London, SE = South East, SW = South West. The 10 LTLAs with highest smoothed prevalence were all in South West: Stroud, Cheltenham, Gloucester, South Gloucestershire, Bath and North East Somerset, Wiltshire, Cotswold, Swindon, Tewkesbury, and City of Bristol

**Supplementary Figure 3.**
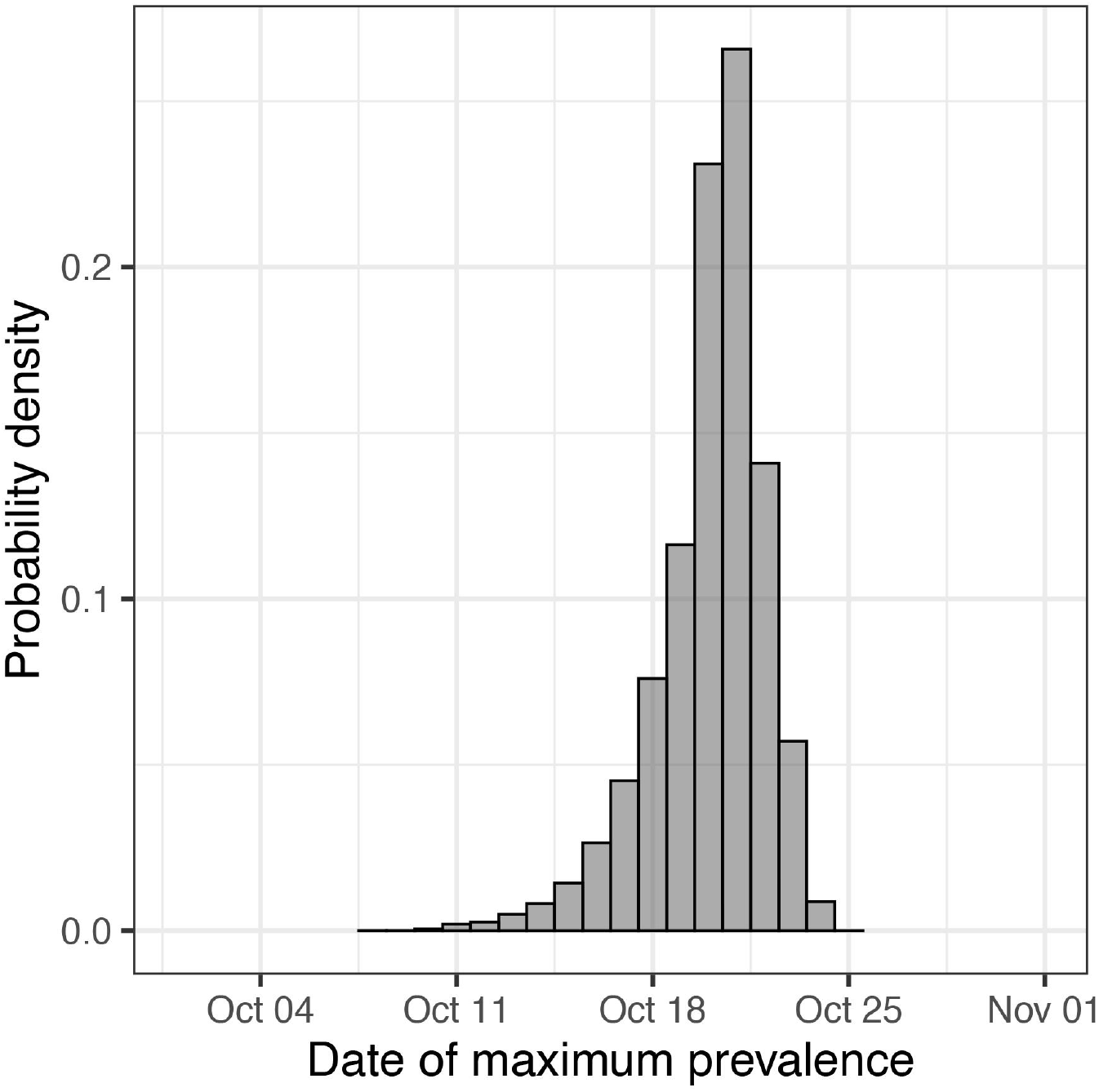
Posterior probability density for the estimated date at which weighted prevalence, as estimated from the P-spline model, was at a maximum during the period of round 14 to round 15.

## Footnotes

1 Vaccination status for linked data was defined using time since last vaccination. Unvaccinated are those not having received any vaccine dose or one dose less than 14 days before swabbing; double dose vaccinated are those having received their second dose 14 days or more before swabbing

2 We considered that the effect of the third dose was effective 14 days or 7 days after vaccination. Participants with two doses are thus defined as those having only received two vaccine doses or three doses with the third one administered less than 14 (or 7) days prior to swabbing; participants with three doses are those who received their third vaccine dose 14 (or 7) days prior to swabbing.

